# Emulating a target trial to assess effect modification: an application to obesity in the comparative effectiveness and safety of apixaban versus warfarin in non-valvular atrial fibrillation using electronic health records

**DOI:** 10.1101/2023.10.12.23296756

**Authors:** Turki M Bin Hammad, Emma Powell, Paris J Baptiste, Ian Douglas, Kevin Wing

## Abstract

Anticoagulation therapy is recommended for patients with non-valvular atrial fibrillation (NVAF) and an increased risk of stroke. Although apixaban showed superiority over warfarin in the Apixaban for Reduction in Stroke and Other Thromboembolic Events in Atrial Fibrillation (ARISTOTLE) study, little is known about their effects in overweight and obese patients, with concerns that obesity might undermine apixaban’s effects due to the fixed dosing across body mass index (BMI) groups. We emulated a target trial similar to the ARISTOTLE study using Clinical Practice Research Datalink (CPRD) linked data to estimate the 36-month risk ratios (RR) and risk differences of the effects of apixaban compared to warfarin in NVAF patients in a composite of stroke/systemic embolism (SE), major bleeding and all-cause mortality. In 55,826 patients, apixaban did not differ across groups of BMI in stroke/SE with RR (95% CI) of 1.15 (0.81, 1.62) in normal weight, 1.06 (0.70, 1.61) in overweight and 1.23 (0.69, 2.17) in obese patients. In major bleeding, apixaban was not superior to warfarin in the normal weight group (RR (95% CI) 1.10 (0.76, 1.60) but superior in overweight (RR (95% CI) 0.73 (0.57, 0.93) and obese (RR (95 % CI) 0.67 (0.52, 0.87) groups. In NVAF, the effectiveness and safety of apixaban compared to warfarin were consistent across BMI groups.

## Introduction

Atrial fibrillation (AF) is a disease of irregular heart rhythm and the most common form of arrhythmia. It affects more than 46 million people around the world increasing the risk of stroke and mortality^1–4^. Patients at increased risk of stroke are recommended to start anticoagulation therapy. Warfarin, a vitamin K antagonist (VKA), was the anticoagulant of choice for a long period before a direct oral anticoagulant (DOAC), apixaban, proved to be superior in reducing the risk of a composite of stroke and systemic embolism and the risk of bleeding in the Apixaban for Reduction in Stroke and Other Thromboembolic Events in Atrial Fibrillation (ARISTOTLE) trial in 2011^5–7^.

Randomised controlled trials (RCTs) are usually conducted at a high cost limiting their representation of important subpopulations^8^. Analysis of electronic health records (EHRs) addresses this issue but comes at the expense of estimating treatments’ effects in the absence of randomisation rendering any observed associations prone to confounding^9^. Statistical techniques (e.g., standardisation or outcome-regression) can only account for known and collected variables while those unmeasured can still cause biased causal treatment estimates^10^.

One way to improve the validity of inferences from observational data is to benchmark or validate an observational study against an existing RCT. That is, to design an observational study that mirrors the reference RCT in terms of design and statistical analysis and then compare the results from the two studies^11,12^. If results are comparable, one can have more trust in this data source to explore treatment effects in under-represented populations. Powell *et al*.(2021,2022) replicated the ARISTOTLE trial using a trial-analogous cohort from the Clinical Practice Research Datalink (CPRD) linked to Hospital Episodes Statistics (HES) and Office for National Statistics (ONS) databases in the United Kingdom^13,14^.

Obesity can play an important role in the treatment response of the two anticoagulants. Warfarin is a highly lipophilic drug with only a small fraction of the total dose unbounded (the active part of the drug) from plasma protein. This is affected by changes triggered by obesity^15–17^. For apixaban, a lower dose is used in underweight patients but there is no dose modification for obese patients possibly leading to sub-optimal dosing. Recent literature identified a gap in the evidence for optimal dosing of patients with increasing body weight which results in altered haemostasis and increased thrombosis^18,19^.

We emulated a target trial similar to ARISTOTLE to investigate whether obesity is an effect modifier of the effectiveness and safety of apixaban compared to warfarin in NVAF patients.

## Methods

### Study design

We carried out a cohort analysis emulating a target trial highly similar to the ARISTOTLE study. We had two primary clinical questions: What is the effect of apixaban (5mg/2.5mg twice daily) compared to warfarin (targeting an INR of (2.0-3.0)) in NVAF patients across each BMI group on the 3-year risk of 1) a composite of ischaemic/haemorrhagic stroke or systemic embolism regardless of treatment discontinuation or switching. 2) bleeding had no patient discontinued or switched treatment.

Our estimands targeted 1) treatment effects of apixaban to warfarin in NVAF patients regardless of treatment discontinuation or switching for each effectiveness outcome (Table S1, Estimands 1,3-7). 2) An estimand that differed in handling intercurrent events of treatment switching and discontinuation by using a hypothetical strategy through censoring for the safety outcome (Table S1, Estimands 2). We considered death as a mediator for other outcomes as our interest was the total effect of treatments^20^ .Figure 1 shows timelines of data collection^21^.

**Figure 1.**
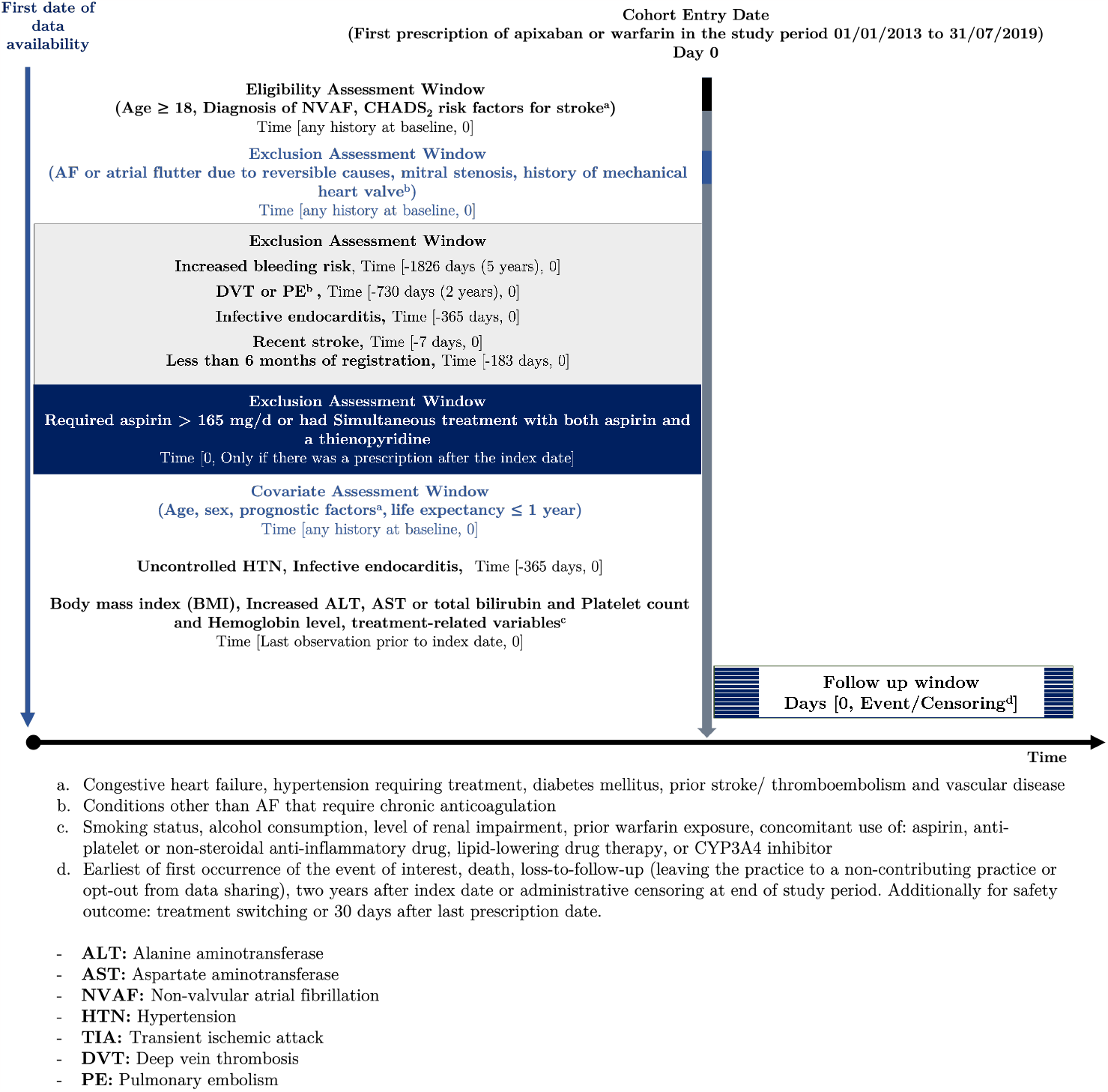
Study scheme and timelines showing windows and time-periods for collecting study variables

#### The reference RCT: ARISTOTLE study^5^

##### Study design

ARISTOTLE was a randomised, double-blind, double-dummy, active-controlled study that compared apixaban (5mg/2.5mg) to warfarin (targeting INR of 2.0 to 3.0) in AF patients from 19/12/2006 to 30/01/2011.

Patients were 18 ≥ years, had permanent or persistent AF or atrial flutter, had at least one additional risk factor for stroke. Inclusion and exclusion criteria are detailed in Table S2.

The primary objective was to establish the non-inferiority of apixaban to warfarin in terms of the primary outcome of a composite of ischaemic/haemorrhagic stroke or systemic embolism.

##### Study results

In 18,201 patients, the hazard ratio (HR) (apixaban/warfarin) for the primary composite outcome was 0.79 (95% CI: 0.66, 0.95; *p*: <0.001 for non-inferiority; *p*: 0.01 for superiority). The HR for major bleeding was 0.69 (95% CI: 0.60, 0.80; *p*: <0.001) and for death from any cause 0.89 (95% CI: 0.80, 0.998; *p*: 0.047).

#### The target trial protocol

Our target trial design consisted of two steps: 1) defining a protocol and an analysis plan of the target trial that would have been conducted if feasible; 2) emulating the target trial using observational analysis of CPRD-Aurum linked data apart from aspects that are limited by the nature of the data^22,23^. Table S2 shows the protocol of the target trial. This would be a pragmatic, open-label, randomized controlled trial closely mirroring the design and analysis of the ARISTOTLE study while considering the settings of the current study.

The main deviations in our target trial: 1) An open-label trial design since it is not feasible to emulate a double-blind trial in routine care^23^ ; 2) Study duration was from 01/01/2013 to 31/07/2019; 3) It included incident users of treatments; 4) Randomization was stratified by BMI to have a more efficient stratified analysis; 5) It did not assess some of the secondary outcomes. A detailed comparison can be found in Table S2.

#### Observational cohort analysis

We emulated the target trial using Clinical Practice Research Datalink Aurum (CPRD-Aurum) linked data with deviations such as the lack of informed consent and timeline of assessing baseline variables.Table S2 highlights the differences between the target trial and the observational analysis.

### Data sources

The CPRD-Aurum contains anonymized primary care EHRs universally accessed across the United Kingdom (UK). It currently covers around ∼20% of the total population, with about 38.4 million patients available for linkage^24^.

The CPRD data is representative of the UK population in terms of age, sex, ethnicity, and BMI^25,26^ with only 5.4% of individuals registered with a general practice opting-out of data sharing^27^.

The database contains information on demographic characteristics, diagnoses and symptoms, drug prescriptions, vaccination history, laboratory tests, referrals to hospital and specialist care, and lifestyle factors (e.g., alcohol consumption).^28^.

The database was linked to the index of multiple deprivation, a measure of socioeconomic status in the UK^29^.

The database is also linked to Hospital Episodes Statistics-Admitted Patient Care (HES-APC) and mortality data from the Office for National Statistics (ONS)^30,31^.

### Setting and participants

We included adult (≥ 18 years old) patients registered for ≥ 6 months in CPRD-Aurum participating general practices (to ensure they are incident users) with available linkage to HES-APC and ONS. The study was restricted to patients residing in England because of the availability of linkage to HES-APC^32^. The study period started on 01/01/2013 (after apixaban gained regulatory approvals) to 31/07/2019. Participants were followed from the date of their first prescription of study treatments to the earliest of first occurrence of study outcomes or censoring events.

Detailed eligibility criteria can be found in Table S2. Participants with a first prescription of apixaban or warfarin during the study period had to have documented NVAF diagnosis and scoring ≥ 2 in CHA_2_DS_2_-VASc. Patients were excluded if they had a history of clinically significant mitral stenosis, increased bleeding risk that contraindicates oral anticoagulation, other conditions requiring chronic anticoagulation, active infective endocarditis, use of aspirin >165 mg/day or simultaneous treatment of both aspirin (any dose) and clopidogrel or ticlopidine, stroke within seven days before the index date.

### Variables

Details of data sources, codes and levels of study variables can be found in Table S3.

#### Treatment Strategies

We compared two treatment strategies 1) apixaban (2.5 mg/5 mg); 2) warfarin targeting an INR level of 2.0 to 3.0. We used the CPRD-Aurum to classify participants to a treatment strategy based on their earliest treatment prescription.

#### Outcomes

The primary outcomes in the study were risk of 1) composite outcome of stroke or SE and 2) major bleeding (defined as haemorrhagic stroke, gastrointestinal, retinal, respiratory and other unspecified sites haemorrhages).

Secondary outcomes were risk of the individual components of stroke/SE and death from any cause.

Study outcomes were identified using HES-APC and ONS based on the International Statistical Classification of Diseases and Related Health Problems version 10 (ICD-10) clinical codes.

Linkage to HES-APC reduces the chance of under-ascertainment of study outcomes because they are usually managed at secondary care points^33,34^.

#### Covariates

We used directed acyclic graphs (DAGs)^35^ to depict our assumptions about the underlying confounding (Figures S1, S2, S3). We assumed that patients were randomly assigned to a strategy conditioning on their baseline covariates and the DAGs were used for confounder selection in our statistical models.

For all outcomes, we accounted for age, gender, ethnicity, individual-level and practice-level indices of multiple deprivation (IMD2015), alcohol, cancer, smoking, renal function, liver disease, persistent uncontrolled hypertension (based on two readings in the prior year), low platelet count (≤ 100,000/mm^3^), hemoglobin level <9 g/dL, concomitant use of low-dose aspirin or clopidogrel, and index year (to account for changes in clinical care over the years).^36–56^

For effectiveness outcomes, we additionally included the remaining CHA_2_DS_2_-VASc score components and concomitant use of medications (statins, beta-blockers, amiodarone, angiotensin-converting enzyme (ACE) inhibitors or angiotensin receptor blockers (ARBs)).^47,57,58,58–62^

For mortality we added the components of the Charlson comorbidity index. These were chronic obstructive pulmonary disease, connective tissue disease, peptic ulcer disease, hemiplegia, solid and blood tumors.^63–65^

For bleeding, we included components of the ORBIT Bleeding Risk Score for Atrial Fibrillation and HAS-BLED Score for Major Bleeding Risk. These factors included any history of GI bleeding, intracranial bleeding, or haemorrhagic stroke, hypertension^66,67^. These were dealt with at the design stage through restriction.

We considered BMI as an effect modifier and was included in the causal graphs^68^. Although obesity can impact treatment response, it is unlikely to affect treatment prescription through paths other than one of the CHA_2_DS_2_-VASc components, which was blocked (Figures S1, S2, S3). BMI was recorded using the latest observation before study entry based on height and weight or direct BMI records and categorised as either healthy weight (≤ 24.9 kg/m^2^), overweight (25-29.9 kg/m^2^) or obese: (≥ 30kg/m^2^).

### Statistical methods

We used a complete case analysis as our analysis population. We used stabilized inverse probability treatment weighting (IPTW) to achieve exchangeability between study arms based on baseline confounders. The weights were estimated using logistic regression models with treatment assignment as the outcome and confounders as predictors. We then calculated the predicted probability of treatment for each person and assigned it as the denominator of the IPT weights. The numerator for IPT weights targeting effect modification was estimated from the predicted probability using a logistic regression model with treatment assignment as the outcome and BMI groups as the only predictor. We also accounted for bias arising from censoring due to loss to follow-up using inverse probability censoring weighting. We obtained the weights using a pooled over time logistic regression model to estimate the time-varying probability of censoring with treatment and the same covariates in the IPT model. To get the final weights, we multiplied the weights from the inverse probability treatment model to those from the censoring model (Section F). We then used the weights in an IPW estimator based on weighted estimates of the observed cause-specific hazards of the outcome and death as a competing event (coincides with a weighted Aalen-Johansen estimator for competing risks) stroke/SE and bleeding outcomes and a complement of a weighted product-limit (Kaplan–Meier) estimator for death to estimate the effects of treatments on the 3-year cumulative incidence of outcome (36 months) with time since first prescription as the time scale and the risk of outcome estimated each month in the study^20^. We reported BMI-stratum-specific risk differences per 100 people, risk ratios and ratios of risk ratios. We derived 95% confidence intervals (CI) using the 2.5 and 97.5 percentiles of a non-parametric bootstrap distribution using 500 samples. We assessed both additive and multiplicative effect modification using the warfarin’s normal weight group as the reference arm but did not test for interaction^69,70^. Participants were censored in case of leaving the practice, opting-out from data sharing, or at the end of the study period.

We conducted one additional analysis using a hypothetical estimand for the primary effectiveness outcome given the potential importance for clinicians (Table S1, Estimands 8). We also conducted four sensitivity analyses: 1) Using 99% truncated IP weights; 2) Restricting the analysis to patients with available BMI measurements in the 3 years prior to study entry 3) Estimating the direct effect using an IPW estimator that corresponds to the complement of a weighted product-limit (Kaplan–Meier) estimator by censoring for death 4) Using IP weighted Cox proportional hazard model.

## Results

### Participants

55,826 (28.8%) out of 194,210 participants with a diagnosis of NVAF and first prescription of apixaban or warfarin between January 1, 2013, to July 31, 2019, were eligible for the study. There were 30,420 apixaban users contributing 43,472 person-years of follow-up and 25,406 warfarin users contributing 57,752 person-years of follow-up. Figure 2 shows the flow of participants selection in the study.

**Figure 2.**
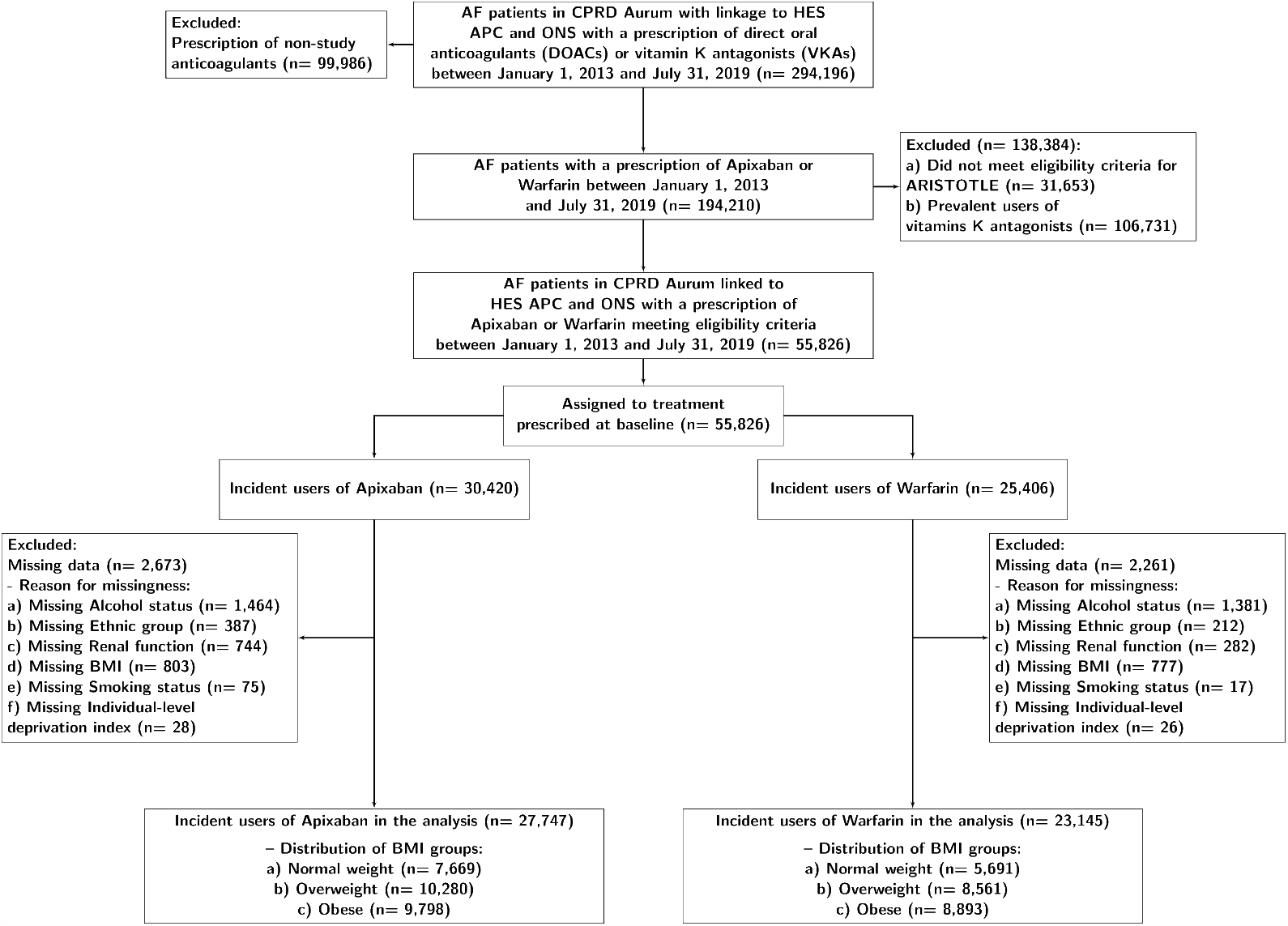
Flowchart of eligibility of patients diagnosed with non-valvular atrial fibrillation and had a first prescription of an anticoagulant during study period

Table 1 shows the baseline characteristics of the study population by exposure. In general, the characteristics were well-balanced between the two treatment arms. There were, however, noted differences in terms of some prognostic factors. Apixaban was given more frequently to patients with a history of stroke, transient ischemic attack, or systemic embolism (27% compared to 21%) but less frequently in other vascular diseases compared to warfarin. Standardised mean differences in baseline characteristics were highly balanced after IPT weighting Table S4. A summary of IP weights can be found in Section F.

**Table 1.**
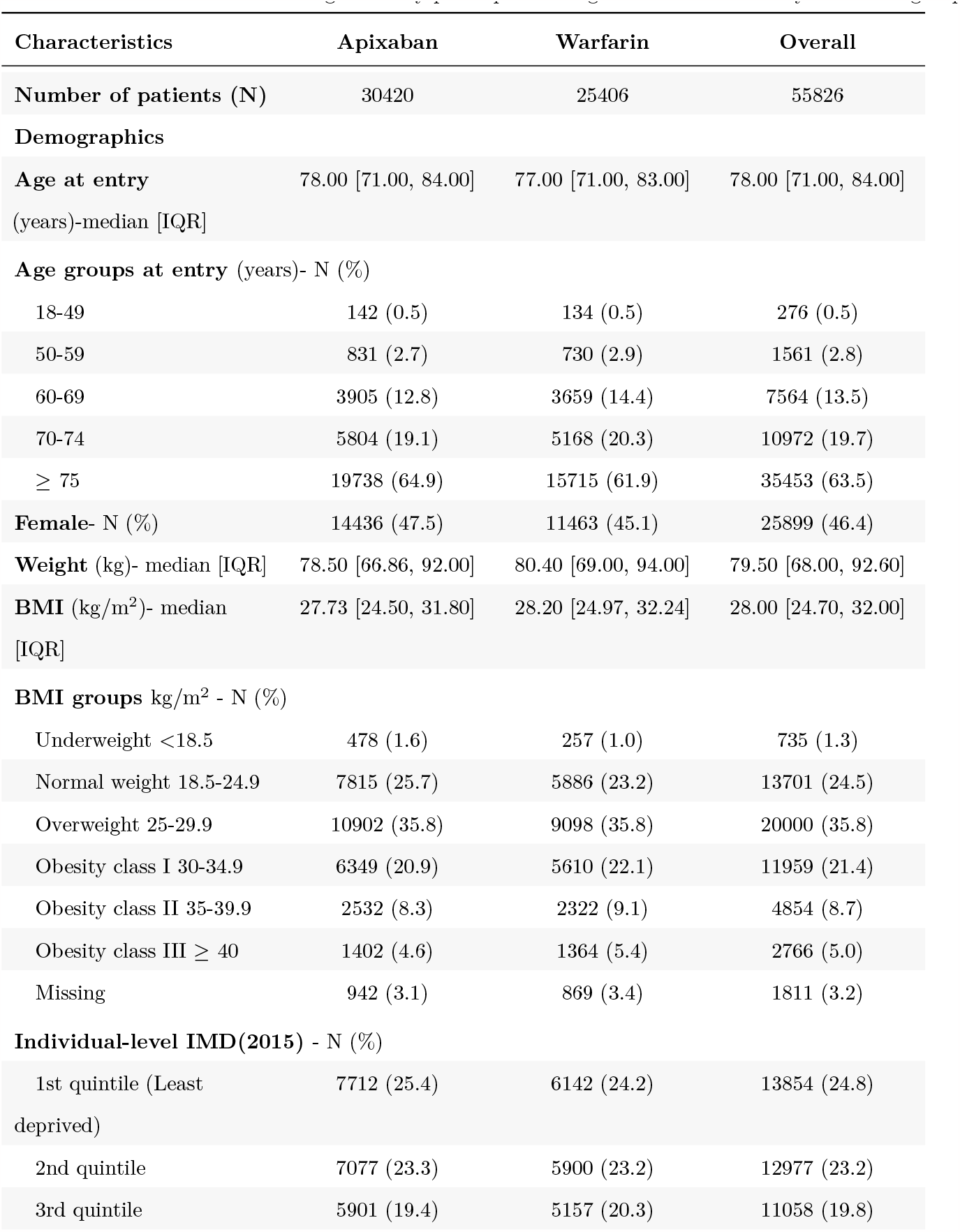
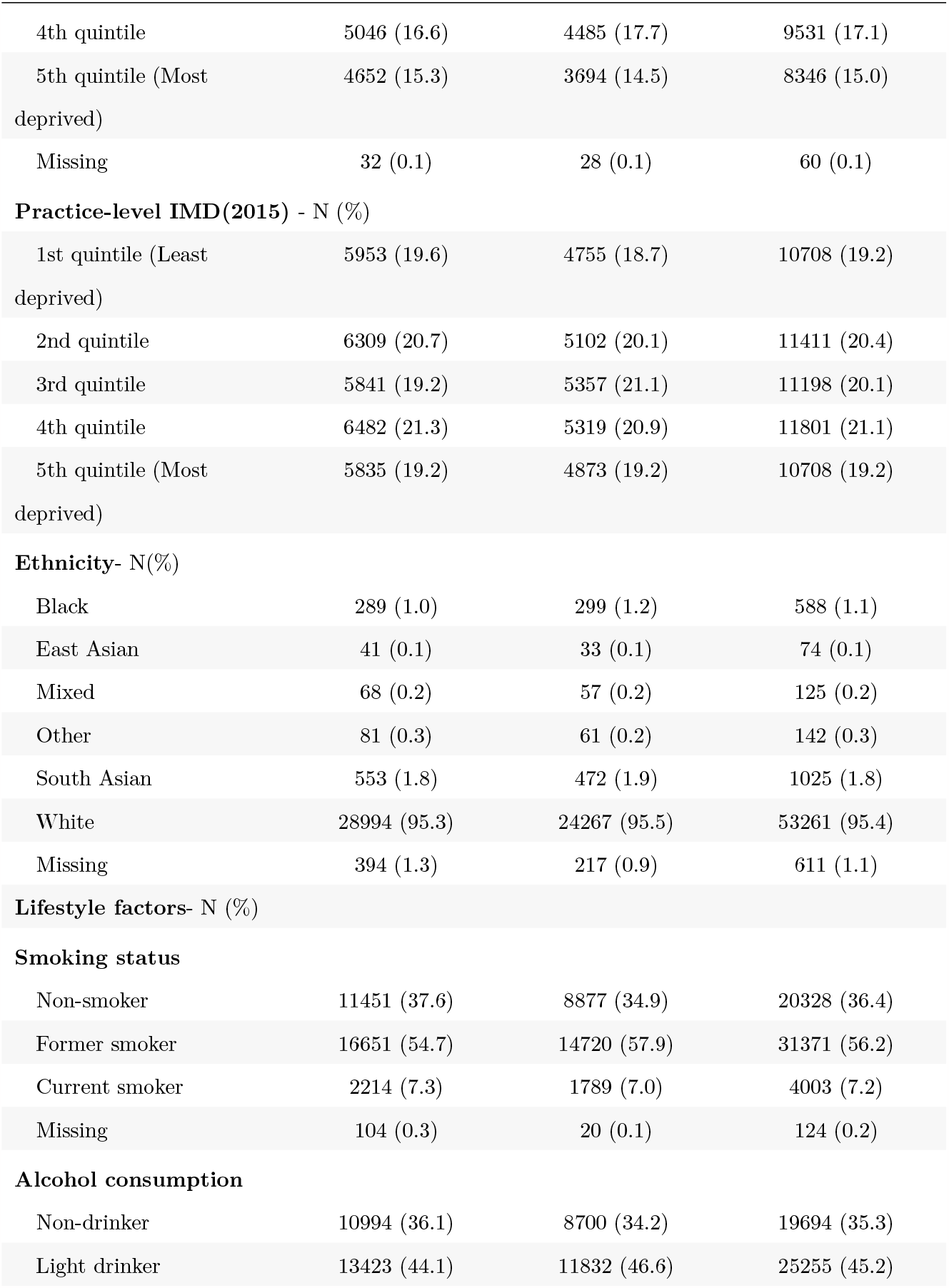
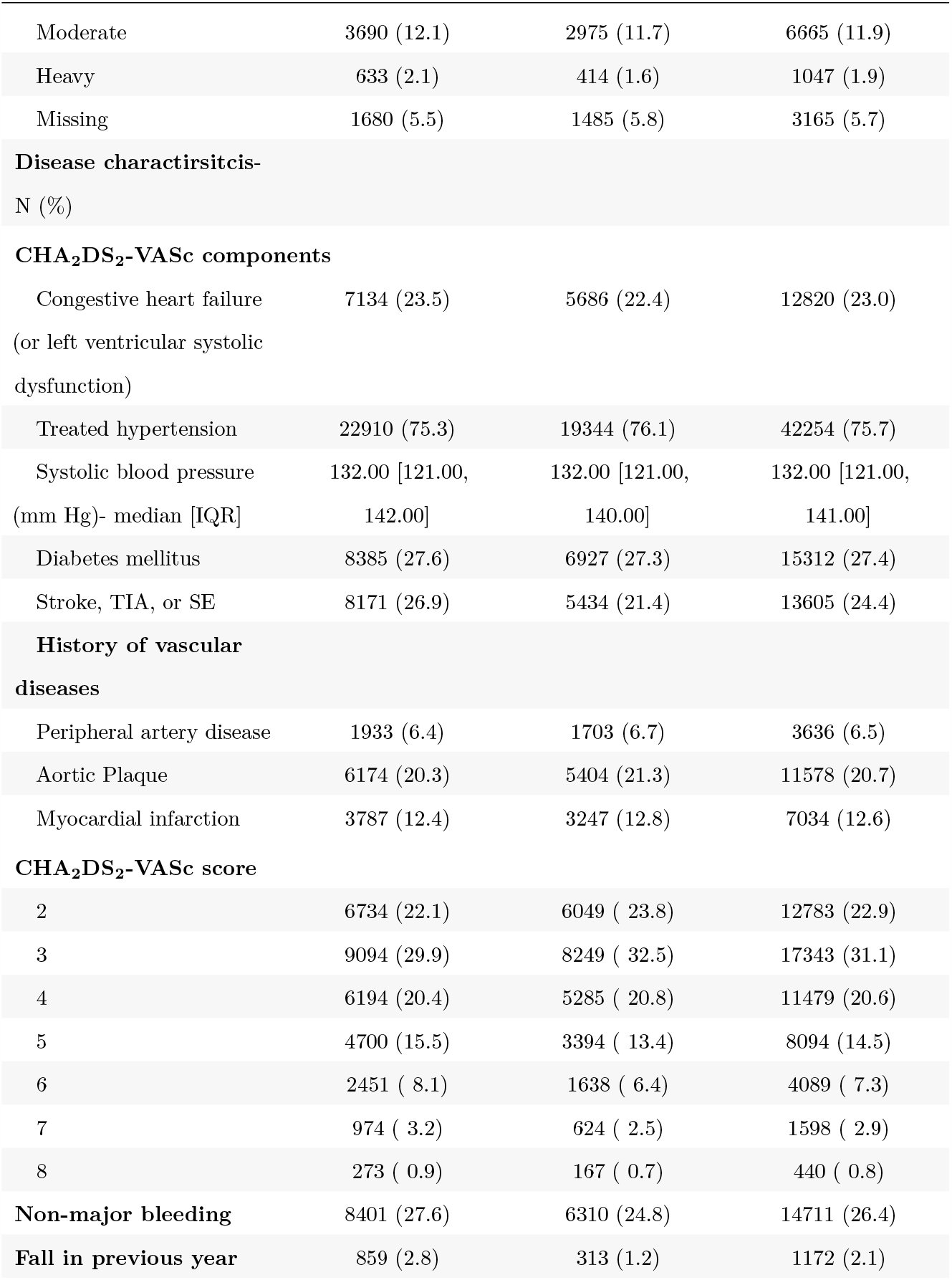
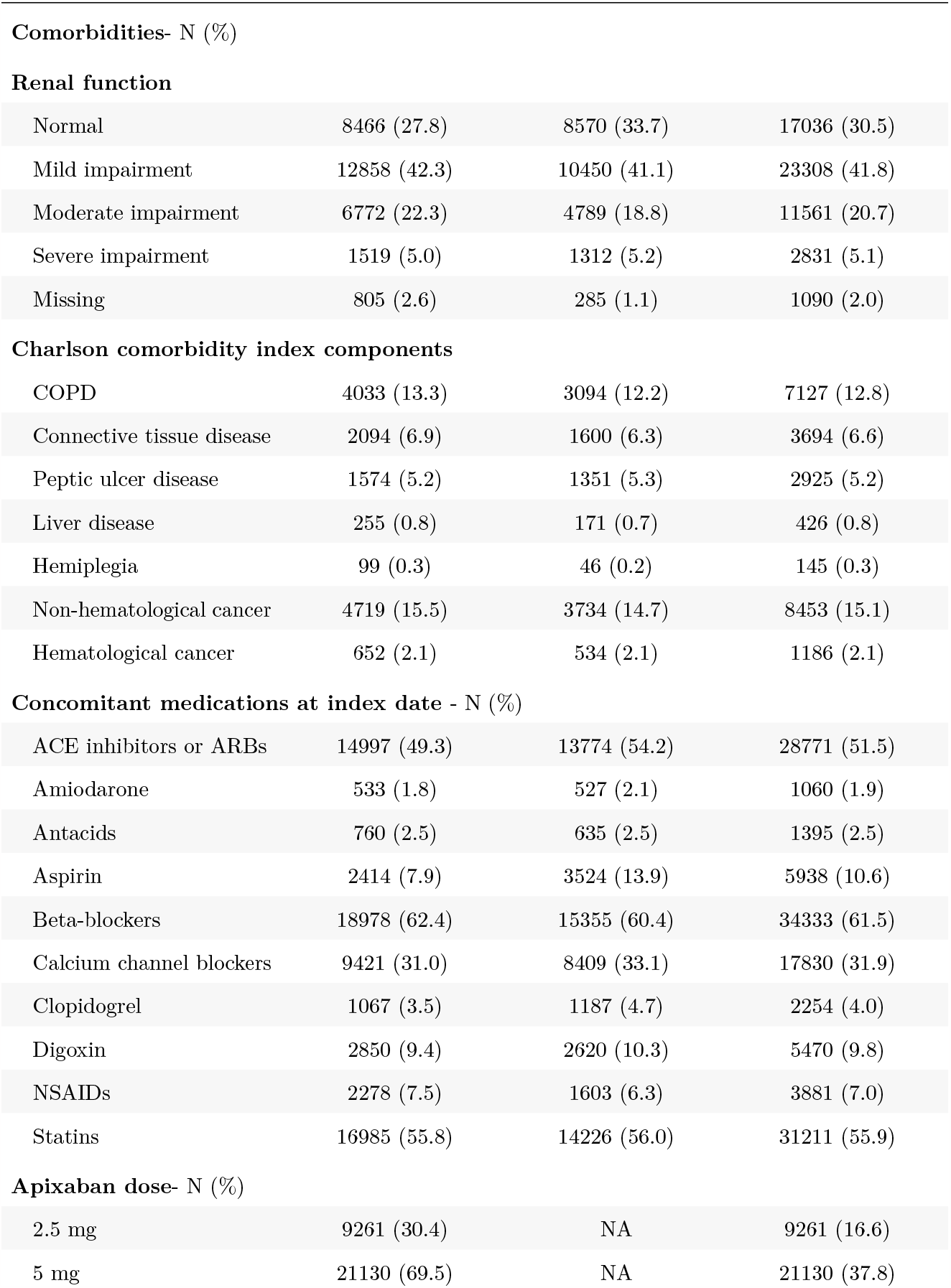
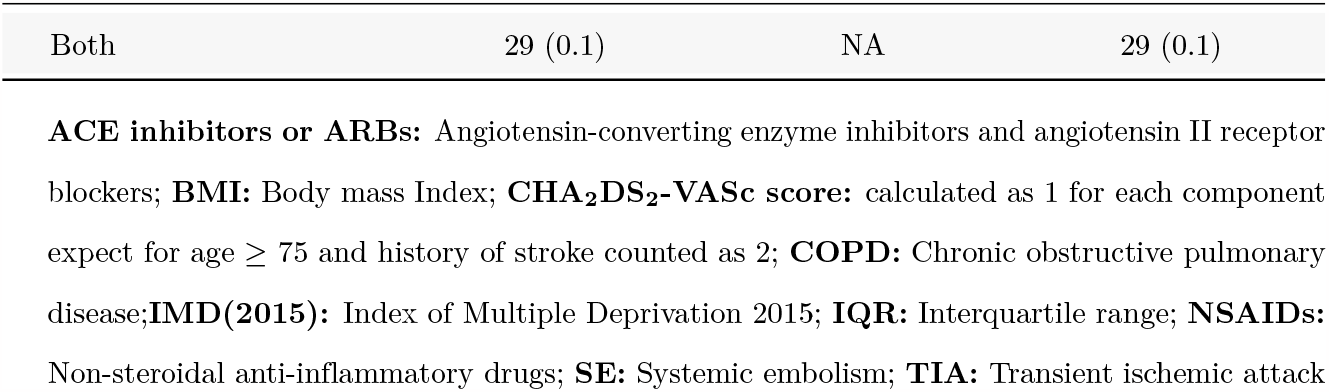
Baseline characteristics of eligible study participants using CPRD linked data by treatment group.

### Comparative effectiveness

#### Primary outcomes

##### Stroke/systemic embolism

For the primary composite endpoint, the risk ratio (95% CI) of apixaban to warfarin was 1.15 (0.81, 1.62) in the normal weight group, 1.06 (0.70, 1.61) in the overweight group and 1.23 (0.69, 2.17) in the obese group. The estimated risk difference per 100 people (95% CI) was 0.6 (−0.7,1.9) in the normal weight group, 0.2 (−1.6, 2.0) in the overweight group and 0.7 (−1.7, 3.2) in the obese group. Figures (3, 4) show the 3-year BMI stratum-specific risks, risk differences per 100 people, and risk ratios and the associated cumulative incidences stratified by BMI. The stroke (any type), ischemic stroke, and SE outcomes were consistent in showing no difference between apixaban and warfarin. In haemorrhagic stroke, apixaban was better than warfarin in overweight patients but failed to show superiority in normal weight and obese patients. All of the individual components were consistent with the composite endpoint in terms of effect modification Section G.

**Figure 3.**
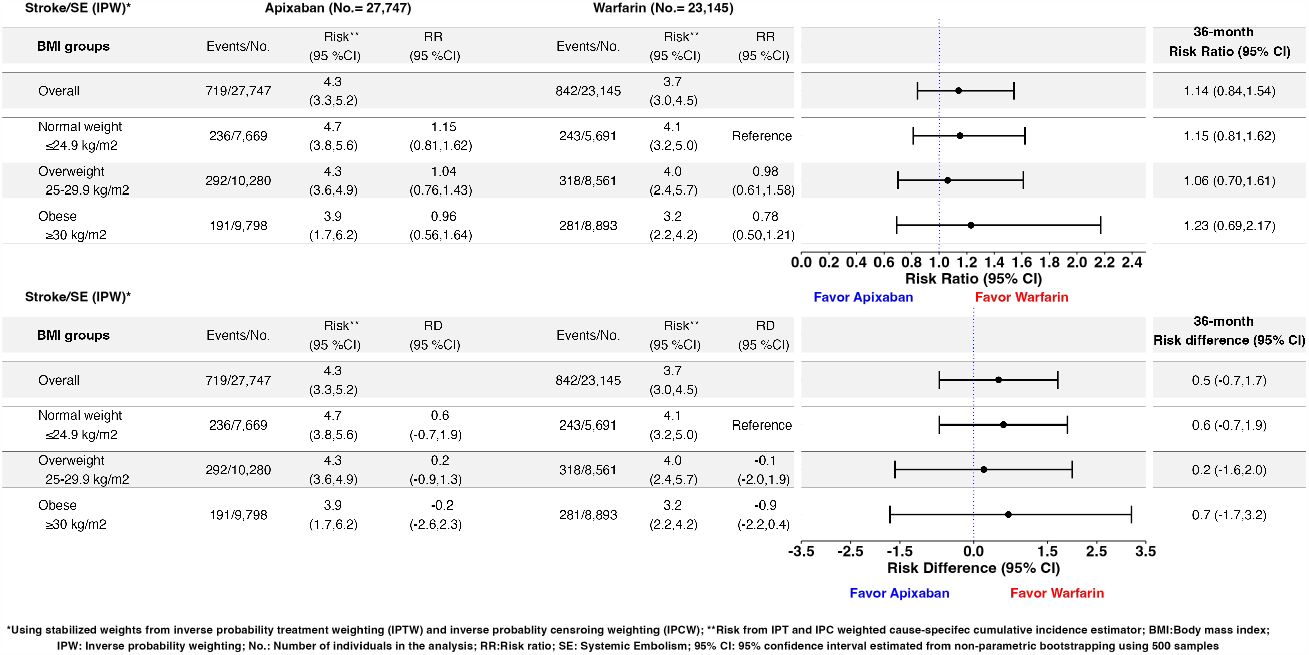
Inverse probability weighted estimates of the 3-year BMI stratum-specific risks, risk differences per 100 people, and risk ratios of the comparative effectiveness for the total effect of apixaban versus warfarin in stroke/systemic embolism using a complete-case analysis (Estimand 1)

**Figure 4.**
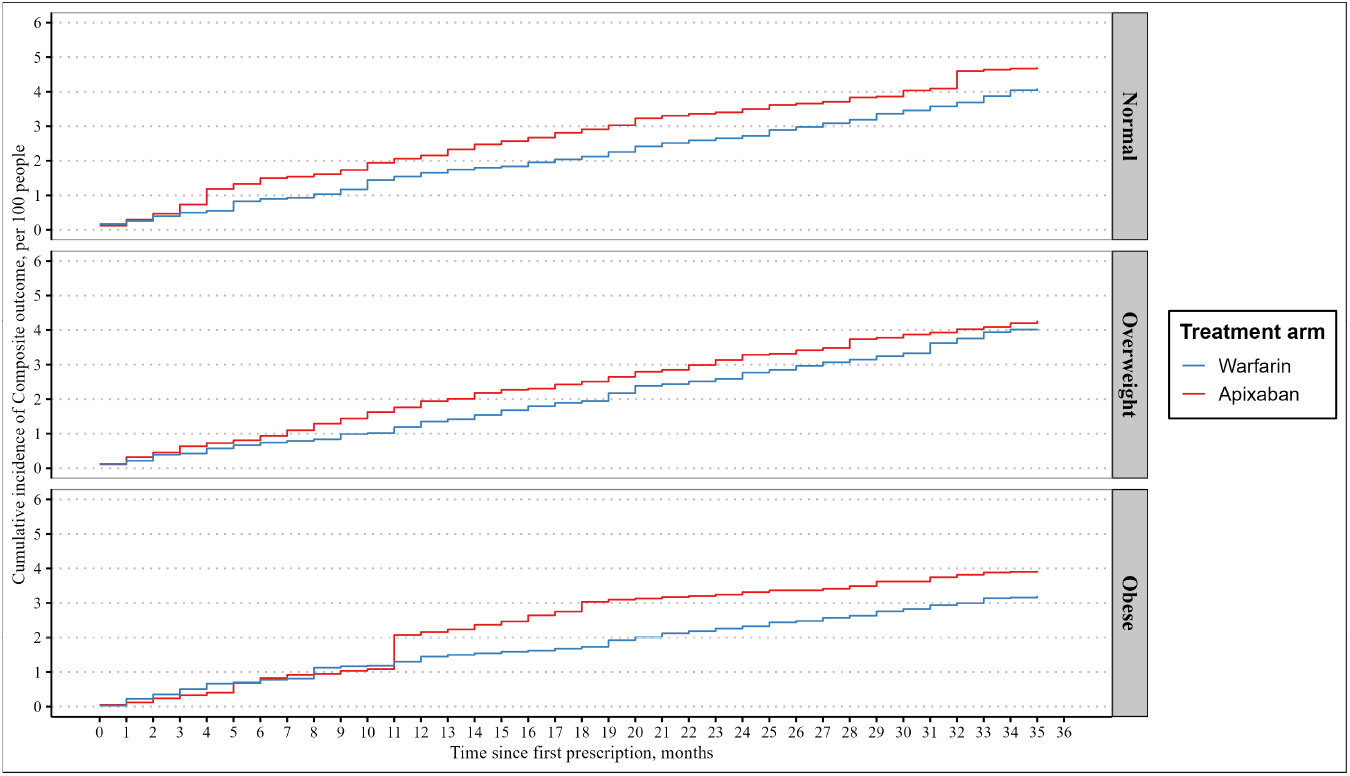
Inverse probability weighted cumulative incidence of the 3-year risk of the total effect of apixaban versus warfarin in stroke/systemic embolism stratified by BMI using a complete-case analysis (Estimand 1)

For death from any cause, the risk ratio (95% CI) of apixaban to warfarin was 1.14 (0.82, 1.60) in the normal weight group, 0.87 (0.61, 1.24) in the overweight group and 1.05 (0.74, 1.50) in the obese group. The estimated risk difference per 100 people (95% CI) was 3.2 (−4.6, 11.0) in the normal weight group, -2.1 (−8.2, 3.9) in the overweight group and 0.8 (−5.1, 6.6) in the obese group Figures (5, 6).

**Figure 5.**
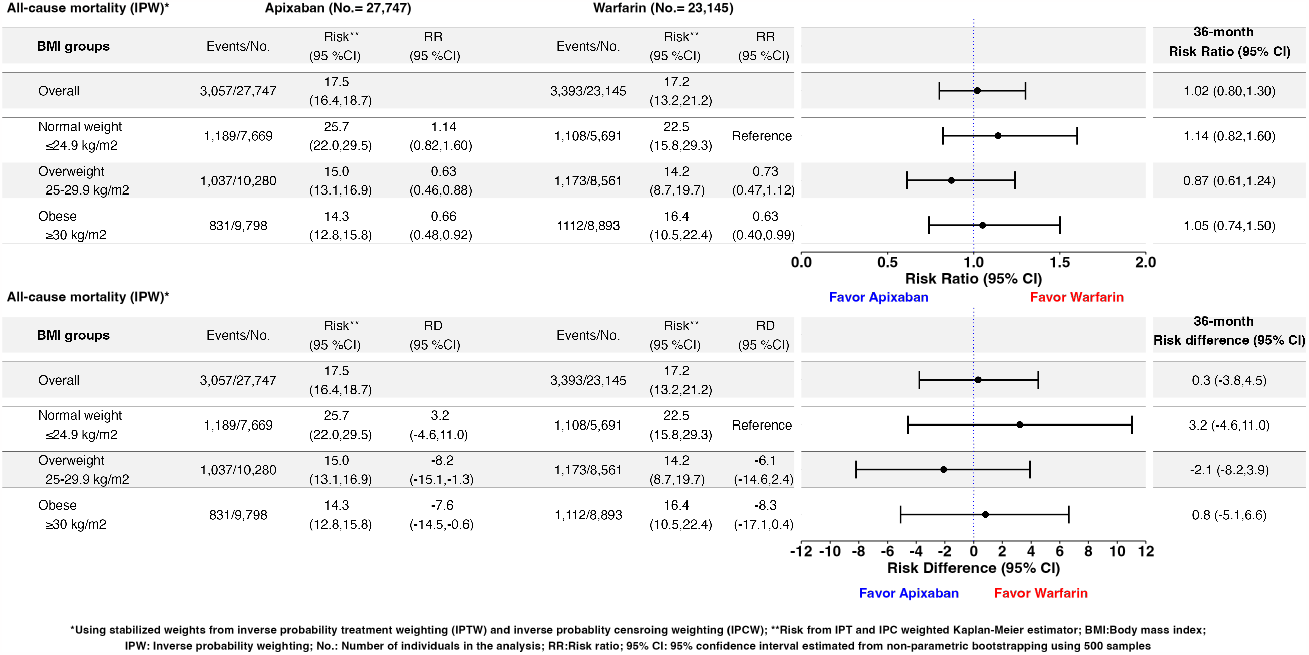
Inverse probability weighted estimates of 3-year BMI stratum-specific risks, risk differences per 100 people, and risk ratios of the comparative effectiveness for the total effect of apixaban versus warfarin in all-cause mortality using a complete-case analysis (Estimand 7)

**Figure 6.**
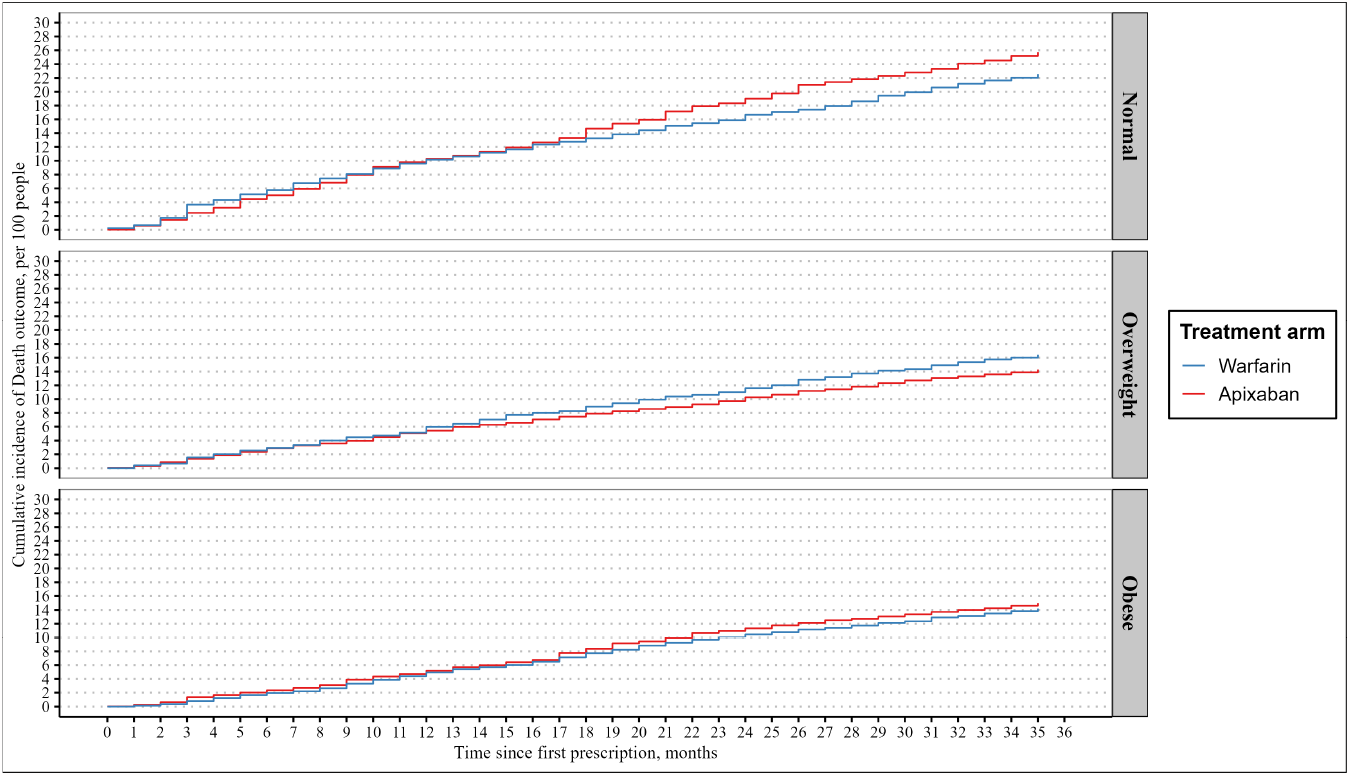
Inverse probability weighted cumulative incidence of the 3-year risk of the total effect of apixaban versus warfarin in all-cause mortality stratified by BMI using a complete-case analysis (Estimand 7)

#### Major bleeding

The risk ratio (95% CI) of apixaban to warfarin was 1.10 (0.76, 1.60) in the normal weight group, 0.73 (0.57, 0.93) in the overweight group and 0.67 (0.52, 0.87) in the obese group. The estimated risk difference per 100 people (95% CI) was 0.9 (−2.4, 4.2) in the normal weight group, -2.3 (−4.1, -0.5) in the overweight group and -3.2 (−5.7, -0.8) in the obese group Figures (7, 8).

**Figure 7.**
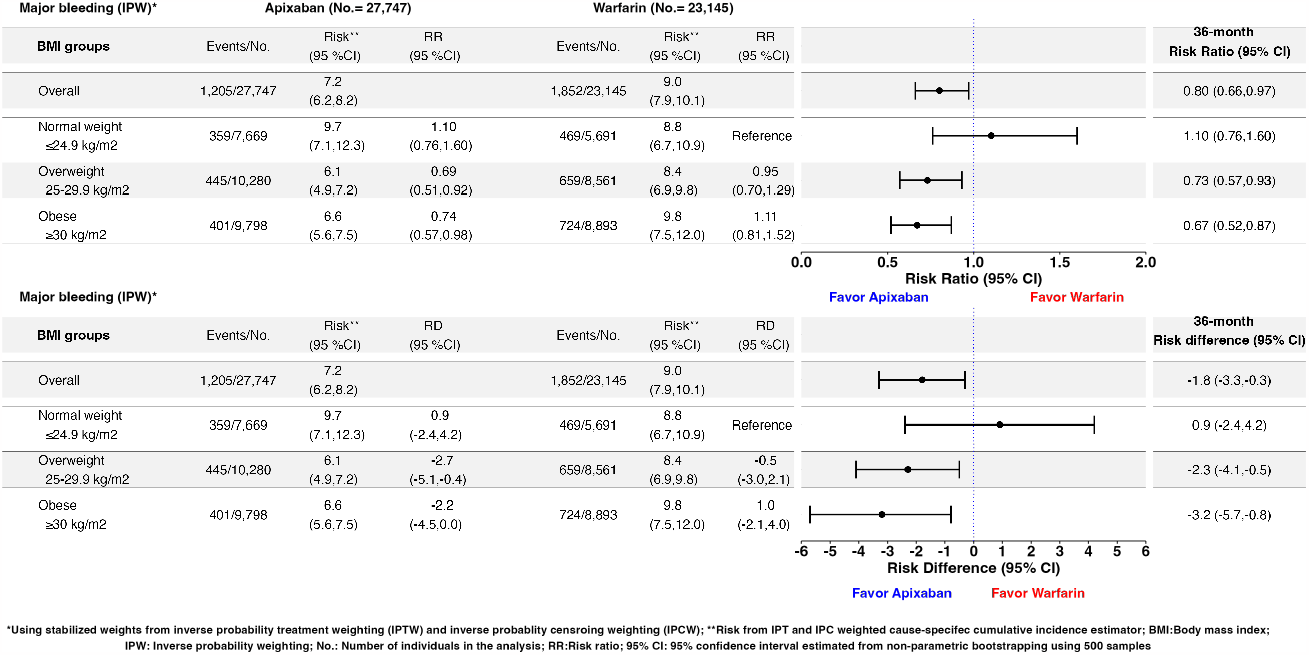
Inverse probability weighted estimates of 3-year BMI stratum-specific risks, risk differences per 100 people, and risk ratios of the comparative safety for the total effect of apixaban versus warfarin in major bleeding using a complete-case analysis (Estimand 2)

**Figure 8.**
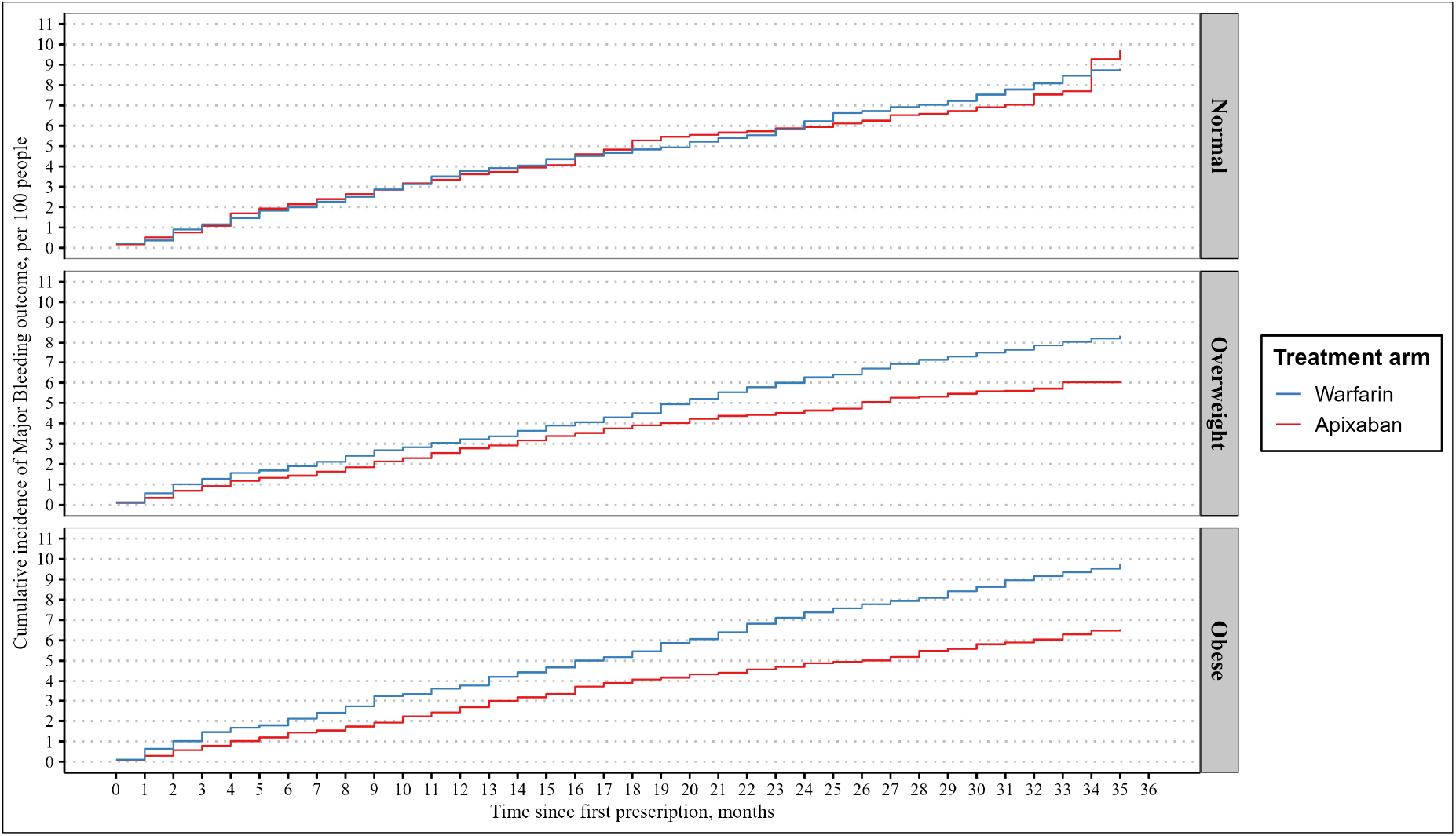
Inverse probability weighted cumulative incidence of the 3-year risk of the total effect of apixaban versus warfarin in major bleeding stratified by BMI using a complete-case analysis (Estimand 2)

#### Supplementary and sensitivity analyses

The hypothetical estimand for primary effectiveness outcome is reported in Figure S9. Sensitivity analyses were overall consistent with the main results (Figures S10, S11, S12, S13, S14, S15 and Section H.4).

## Discussion

In this large cohort study using observational data from the CPRD-Aurum linked to HES APC and ONS to emulate a target trial, we did not find evidence of effect modification by obesity, using BMI as a proxy, on the effectiveness and safety of apixaban compared to warfarin in non-valvular atrial fibrillation patients.

In the analysis, apixaban was not superior to warfarin in reducing the risk of stroke/SE or all-cause mortality in the overall and across any BMI category. For major bleeding, apixaban showed superiority over warfarin overall and in overweight and obese patients but not in patients with normal weight. Apixaban was also not superior in reducing stroke (any type) or ischemic stroke. For haemorrhagic stroke, apixaban was not superior over warfarin except for obese patients but the study was not powered to answer this question. Events of SE were scarce, resulting in very wide 95% CIs and hindering any meaningful interpretations of the results. The results were robust in sensitivity analyses as none meaningfully deviated from the main analysis.

The similar comparative treatment response across BMI groups in the study was contrary to previous suggestions in the literature that obesity plays a major role in treatment effects of anticoagulants^18,19^.

### Interpretation

Our BMI-stratified analysis was different to the reported results from the ARISTOTLE trial, which investigated the effects in the overall groups rather than within BMI groups^5^. In ARISTOTLE, apixaban was found to be superior to warfarin in both composite of stroke/SE (HR 0.79; 95% CI: 0.66, 0.95) and major bleeding (HR 0.69; 95% CI: 0.60, 0.80). However, our findings were consistent with the subgroup analysis of EU randomised patients of ARISTOTLE showing an HR of 0.92 (95% CI: 0.56,1.52) for stroke/SE^71^. Our findings were consistent with the results of the recent study by Deitelzweig *et al*.(2022) in the United States, which investigated the effects across BMI groups and showed similar conclusions to our study^72^. Another US-based study by Deitelzweig *et al*.(2020) investigated the effects in the obese population only and had similar results to the obese group in our study^73^.

In our study, and similarly, in both US-based cohort studies, the main difference from ARISTOTLE was the lack of superiority for the primary effectiveness outcome (stroke/SE) which could have arisen due to multiple reasons. First, we cannot rule out the possibility of unmeasured confounding. Despite accounting for several confounders in the analysis, there could be psychological and personal factors affecting the choice of prescribed treatment and the risk of the outcome. As the results of ARISTOTLE were published before our study start date and clinical trials usually change the clinical practice, it is possible that treatment choice relied on non-measurable behaviors (e.g., more willingness to comply with INR monitoring despite the availability of other effective treatments).

Second, this could be due to better warfarin control since the median percentage of time in therapeutic range (TTR) (INR 2-3) was 66% for the trial population compared to 76% in the CPRD cohort^74^.

Third, this could be due to deviations from ARISTOTLE protocol, such as lack of blinding or differences in warfarin monitoring. In ARISTOTLE, nonetheless, apixaban was not superior to warfarin in ischemic stroke which comprised a higher proportion of stroke events, in each BMI group, compared to ARISTOTLE. This could have driven the composite endpoint towards the null.

In terms of major bleeding, our study agreed with previous findings for overweight and obese groups but not for normal weight group. Apixaban appeared to be superior to warfarin in major bleeding events, with the effect being consistent across BMI groups. The rate of major bleeding was similar to that reported in ARISTOTLE.

Our study also employed a slightly different causal contrast compared to the other studies. This study was mainly concerned with the effect of prescribing either apixaban or warfarin on the rates of Stroke/SE or bleeding regardless of treatment switching except for safety.

### Strengths and limitations

We used a validated and representative routinely collected primary care data with linkage to secondary care, which improved the completeness of ascertaining study outcomes. We designed the study to emulate a target trial and restricted the enrollment to incident users to minimize bias. Our study also collected and accounted for multiple confounding factors, including demographic, prognostic, social, lifestyle, and socioeconomic factors, including both individual level and practice level deprivation indices. We included a study population similar to those trial-eligible allowing a better comparison with the reference RCT.

Moreover, another strength was the ability to look at BMI groups with acceptable precision. The large sample size (> 50,000 participants) could be looked at from two perspectives. Although this is not largely distinguished in the literature, this study focused on effect modification rather than interaction^70^. That is, what is the average treatment effect within BMI strata rather than, for example, what is the joint effect of anticoagulation and weight loss (i.e., interaction effect). If one would consider the first case, the study was well-powered. However, to detect an interaction effect, a much larger sample size is needed, which would, probably, render our study underpowered^75^.

However, our study had some limitations. We had a large proportion of patients being censored with incomplete follow-up. This resulted in more warfarin patients having a complete follow-up compared to those on apixaban leading to differential follow-up, which can be a source of selection bias in the study^22^. A more comparable approach would have been to split the study into two periods: a recruitment period followed by a follow-up period to allow time for follow-up of the last recruited patient, as in RCTs. This was not feasible in our study since apixaban prescription increased dramatically during the last three years of the study^6^.

Furthermore, we reported both treatment policy and hypothetical estimands but did not measure treatment compliance, which can affect study estimates^76^. Lower compliance can attenuate differences between treatments or, in extreme cases, favor one treatment over another. It is more sensible to assume that compliance would be mainly affected in the warfarin arm due to the need of continuous monitoring, which, if true, would make warfarin effects underestimated. The effects in this study can be considered the effect of prescribing apixaban or warfarin at baseline rather than the effect of treatment^22,76^.

The effect modifier, BMI, can be subjected to measurement error with the categorization of BMI introducing further misclassification^77^. Although we have repeated the analysis restricting BMI observations to ≤ 3 years without a meaningful difference, BMI is expected to be a time-varying effect modifier that can change with time. Especially if this coincides with a diagnosis of NVAF, for which increased body weight is a major risk factor to sustain^78^. We excluded patients with missing data when estimating the inverse probability weights. This might impacted our study estimates despite the data were likely missing at random given the routine clinical care setting.

## Conclusion

We believe that the results provide evidence of comparative treatment effects between apixaban and warfarin in the investigated BMI groups. Furthermore, our results were similar to the other studies investigating either the overall or BMI-stratum-specific effects of treatments. This provides robust data for evidence-based decisions for both patients and health practitioners regarding treatment choice. This could be more valuable also in times of pandemics where many patients might not be able to maintain the continuous monitoring for warfarin due to their frailty as NVAF patients or where many under-resourced healthcare systems might not have the capability for it.

In conclusion, after emulating the target trial, BMI, as a proxy for obesity, was not an effect modifier of the effectiveness and safety of apixaban compared to warfarin. The effects of the two treatments in the study were similar in obese and overweight patients to normal weight patients across all study outcomes. This provides reassurance about the use of apixaban in these patient populations. However, future research is needed to investigate the generalisability of the results, compare the different available direct oral anticoagulants (DOACs), and assess the effect in extremely obese patients.

## Data Availability

Anonymised data from the Clinical Practice Research Datalink (CPRD), Hospital Episodes Statistics-Admitted Patient Care (HES-APC) and mortality data from the Office for National Statistics (ONS) were obtained after ethical approval and cannot be made publicly available.

## Acknowledgements

Author affiliations: Saudi Food and Drug Authority, Riyadh, Saudi Arabia (Turki Bin Hammad); London School of Hygiene and Tropical Medicine, London, United Kingdom (Turki Bin Hammad, Emma Powell,, Ian Douglas, Kevin Wing); Queen Mary University of London, London, United Kingdom (Paris J Baptiste).

This work was partly supported by the Saudi Food and Drug Authority (SFDA) as a part of a Master of Science (MSc) project for Turki Bin Hammad. The SFDA had no role in the design, review or writing of the manuscript.

The views expressed in this paper are those of the author and not do not necessarily reflect those of the SFDA or its stakeholders. Guaranteeing the accuracy and the validity of the data is a sole responsibility of the research team.

Anonymised data from the Clinical Practice Research Datalink (CPRD), Hospital Episodes Statistics-Admitted Patient Care (HES-APC) and mortality data from the Office for National Statistics (ONS) were obtained after ethical approval and cannot be made publicly available. All diagnostic codes can be accessed from https://datacompass.lshtm.ac.uk/id/eprint/3590/. Analysis code is available at: https://github.com/turkimo/apixaban_warfarin_bmi.

The study was approved by the MSc Research Ethics Committee (Ethics Ref: 27537) at the London School of Hygiene & Tropical Medicine and the Independent Scientific Advisory Committee for MHRA Database Research (approved amendment to 19_066).

