## Supplementary Material for "Emulating a target trial to assess effect modification: an application to obesity in the comparative effectiveness and safety of apixaban versus warfarin in non-valvular atrial fibrillation using electronic health records"

### Appendicies

### A Estimands

**Table S1.** Estimands of the observational study emulating target trial

| Estimand component | Description |
|---|---|
| **Estimand 1** | |
| Population | Adults with non-valvular atrial fibrillation receiving anticoagulants meeting eligibility criteria |
| Variable of interest | Composite of stroke (Ischemic or hemorrhagic) or systemic embolism |
| Treatments | Apixaban (2.5/5 mg) versus Warfarin adjusted to target an INR of (2.0-3.0) |
| Population level summary | BMI stratum specific Risk ratios and Risk differences |
| Intercurrent events (strategies to handle them) | 1- Treatment discontinuation (treatment policy strategy) 2- Treatment switching (treatment policy strategy) |
| **Estimand 2** | |
| Population | Adults with non-valvular atrial fibrillation receiving anticoagulants meeting eligibility criteria |
| Variable of interest | Major bleeding |
| Treatments | Apixaban (2.5/5 mg) versus Warfarin adjusted to target an INR of (2.0-3.0) |
| Population level summary | BMI stratum specific Risk ratios and Risk differences |
| Intercurrent events (strategies to handle them) | 1- Treatment discontinuation (hypothetical strategy) 2- Treatment switching (hypothetical policy strategy) |
| **Estimand 3** | |
| Population | Adults with non-valvular atrial fibrillation receiving anticoagulants meeting eligibility criteria |
| Variable of interest | Ischemic or hemorrhagic stroke |
| Treatments | Apixaban (2.5/5 mg) versus Warfarin adjusted to target an INR of (2.0-3.0) |

| | |
|---|---|
| Population level summary | BMI stratum specific Risk ratios and Risk differences |
| Intercurrent events (strategies to handle them) | 1- Treatment discontinuation (treatment policy strategy)<br>2- Treatment switching (treatment policy strategy) |

**Estimand 4**

| | |
|---|---|
| Population | Adults with non-valvular atrial fibrillation receiving anticoagulants meeting eligibility criteria |
| Variable of interest | Ischemic stroke |
| Treatments | Apixaban (2.5/5 mg) versus Warfarin adjusted to target an INR of (2.0-3.0) |
| Population level summary | BMI stratum specific Risk ratios and Risk differences |
| Intercurrent events (strategies to handle them) | 1- Treatment discontinuation (treatment policy strategy)<br>2- Treatment switching (treatment policy strategy) |

**Estimand 5**

| | |
|---|---|
| Population | Adults with non-valvular atrial fibrillation receiving anticoagulants meeting eligibility criteria |
| Variable of interest | Hemorrhagic stroke |
| Treatments | Apixaban (2.5/5 mg) versus Warfarin adjusted to target an INR of (2.0-3.0) |
| Population level summary | BMI stratum specific Risk ratios and Risk differences |
| Intercurrent events (strategies to handle them) | 1- Treatment discontinuation (treatment policy strategy)<br>2- Treatment switching (treatment policy strategy) |

**Estimand 6**

| | |
|---|---|
| Population | Adults with non-valvular atrial fibrillation receiving anticoagulants meeting eligibility criteria |
| Variable of interest | Systemic embolism |
| Treatments | Apixaban (2.5/5 mg) versus Warfarin adjusted to target an INR of (2.0-3.0) |
| Population level summary | BMI stratum specific Risk ratios and Risk differences |

| | |
|---|---|
| Intercurrent events (strategies to handle them) | 1- Treatment discontinuation (treatment policy strategy) 2- Treatment switching (treatment policy strategy) |

| **Estimand 7** | |
|---|---|
| Population | Adults with non-valvular atrial fibrillation receiving anticoagulants meeting eligibility criteria |
| Variable of interest | Mortality |
| Treatments | Apixaban (2.5/5 mg) versus Warfarin adjusted to target an INR of (2.0-3.0) |
| Population level summary | BMI stratum specific Risk ratios and Risk differences |
| Intercurrent events (strategies to handle them) | 1- Treatment discontinuation (treatment policy strategy) 2- Treatment switching (treatment policy strategy) |

| **Estimand 8** | |
|---|---|
| Population | Adults with non-valvular atrial fibrillation receiving anticoagulants meeting eligibility criteria |
| Variable of interest | Composite of stroke (Ischemic or hemorrhagic) or systemic embolism |
| Treatments | Apixaban (2.5/5 mg) versus Warfarin adjusted to target an INR of (2.0-3.0) |
| Population level summary | BMI stratum specific Risk ratios and Risk differences |
| Intercurrent events (strategies to handle them) | 1- Treatment discontinuation (hypothetical strategy) 2- Treatment switching (hypothetical policy strategy) |

### B Target trial protocol

**Table S2.** Brief protocol of target trial emulation

| Protocol component | ARISTOTLE | Target trial | Observational analysis |
|---|---|---|---|
| **Eligibility criteria** | **Inclusion criteria**<br>- Age $\geq$ 18 y between December 19, 2006, through April 2, 2010<br>- Permanent or persistent AF or atrial flutter on ECG at enrollment; or AF or atrial flutter documented by ECG or as an episode $\geq$ 1 min on rhythm strip, Holter monitor, or intracardiac recording on 2 separate occasions at least 2 week apart in 12 months before enrollment<br>- One or more of the following risk factors for stroke:<br>Age $\geq$ 75 y<br>Prior stroke, TIA, or systemic embolus<br>Symptomatic CHF within 3 mo or LV dysfunction with LVEF $\leq$ 40% by echocardiography, radionuclide study, or contrast angiography<br>Diabetes mellitus<br>Hypertension requiring pharmacologic treatment | - Same as ARISTOTLE apart from:<br>- Study period from 01/01/2013 to 31/07/2019<br>- Only in patients from England with EHR in CPRD Aurum and linkage to HES and ONS linked patients.<br>- Registered with a practice contributing research quality data for at least 6 months on or prior to the index date.<br>- Permanent or persistent AF documentation using ECG is not assessed<br>- Three additional stroke risk factors: Age 64-75, female and vascular disease (i.e. $CHA_2DS_2$-VASc instead of CHADS 2 ) | - Same as target trial apart from:<br>- No informed consent is required but patients might opt-out of data sharing |

*(continued)*

| Protocol component | ARISTOTLE | Target trial | Observational analysis |
|---|---|---|---|
| | - Women of childbearing potential must use contraception to avoid pregnancy during treatment period or for 2 wk after last dose of study medication, whichever is longer<br>- All subjects must provide signed written informed consent<br>**- Exclusion criteria**<br>- AF or atrial flutter due to reversible causes<br>- Clinically significant (moderate or severe) mitral stenosis<br>- Increased bleeding risk believed to be a contraindication to oral anticoagulation (eg, previous intracranial hemorrhage)<br>- Conditions other than AF that require chronic anticoagulation (eg, prosthetic mechanical heart valve)<br>- Persistent uncontrolled hypertension (SBP 180mmHg or DBP 100mmHg)<br>- Active infective endocarditis | - No contraception is required from women of childbearing age but any women with history of breastfeeding or pregnancy in last two years prior to index date was excluded<br>- No assessment of planned procedure<br>- No exclusion for: - Patients with presesitant uncontrolled hypertension<br>- Severe comorbid condition with life expectancy $\leq$ 1 year<br>- Active alcohol or drug abuse or psychosocial reasons that make study participation impractical<br>- No assessment of planned procedure<br>- No exclusion for: - Patients with presesitant uncontrolled hypertension | |

*(continued)*

| **Protocol component** | **ARISTOTLE** | **Target trial** | **Observational analysis** |
|---|---|---|---|
| | - Planned major surgery<br>- Planned AF or atrial flutter ablation procedure<br>- Use of unapproved investigational drug or device in past 30 days<br>- Required aspirin 165 mg/d<br>- Simultaneous treatment with both aspirin and a thienopyridine (eg, clopidogrel, ticlopidine)<br>- Severe comorbid condition with life expectancy $\leq$ 1 year<br>- Active alcohol or drug abuse or psychosocial reasons that make study participation impractical<br>- Recent stroke (within 7 days)<br>- Severe renal insufficiency (serum creatinine level 2.5 mg/dL or calculated creatinine clearance <25 mL/min)<br>- ALT or AST 2 $\times$ ULN or a total bilirubin $\geq$ 1.5 $\times$ ULN (unless an alternative causative factor [eg, Gilbert's syndrome] is identified)<br>- Platelet count $\leq$ 100,000/mm$^3$<br>- Hemoglobin level <9 g/dL<br>- Inability to comply with INR monitoring | - Severe comorbid condition with life expectancy $\leq$ 1 year<br>- Active alcohol or drug abuse or psychosocial reasons that make study participation impractical<br>- Severe renal insufficiency (serum creatinine level 2.5 mg/dL or calculated creatinine clearance <25 mL/min)<br>- ALT or AST 2 $\times$ ULN or a total bilirubin $\geq$ 1.5 $\times$ ULN (unless an alternative causative factor [eg, Gilbert's syndrome] is identified)<br>- Platelet count $\leq$ 100,000/mm$^3$<br>- Hemoglobin level <9 g/dL<br>- No assessment of use of unapproved investigational drug or device in past 30 days<br>Any history of symptomatic CHF or LV dysfunction<br>- No assessment of inability to comply with INR monitoring | |

*(continued)*

| Protocol component | ARISTOTLE | Target trial | Observational analysis |
|---|---|---|---|
| **Treatment strategies** | 1- Starting Apixaban or matching placebo twice daily, with apixaban given in 5-mg doses; 2.5-mg doses were used in a subset of patients with two or more of the following criteria:<br>- an age of at least 80 years,<br>- a body weight of no more than 60 kg, or<br>- a serum creatinine level of 1.5 mg per deciliter (133 $\mu mol$ per liter) or more. | 1- Same as ARISTOTLE | Same as target trial |
| | 2- Warfarin (or matching placebo) as 2-mg tablets adjusted to achieve a target international normalized ratio (INR) of 2.0 to 3.0. Patients who are already receiving a vitamin K antagonist before randomization were instructed to discontinue the drug 3 days before randomization, and the study drug was initiated when the INR was less than 2.0. | 2- Warfarin tablets adjusted to achieve a target international normalized ratio (INR) of 2.0 to 3.0. – Did not include patients already reciving vitamin K antagonists so no washout period | Same as target trial |

*(continued)*

| Protocol component | ARISTOTLE | Target trial | Observational analysis |
|---|---|---|---|
| **Assignment procedure** | Participants will be randomized to a treatment strategy while not being aware of the assigned strategy | Participants will be randomized to a treatment strategy and are aware of the assigned strategy | Participants' strategies were classified according to the observed data at baseline while adjusting for the confounders using inverse probability treatment weighting to emulate randomization |

*(continued)*

| Protocol component | ARISTOTLE | Target trial | Observational analysis |
|---|---|---|---|
| **Outcomes** | Efficacy outcomes:<br>The primary efficacy outcome is:<br>- Time to first occurrence of stroke (ischemic or haemorrhagic) or systemic embolism.<br>The key secondary efficacy outcomes:<br>- death from any cause.<br>- rate of myocardial infarction<br>Safety outcomes:<br>- Major bleeding,<br>- Composite of major bleeding and clinically relevant nonmajor bleeding.<br>- Any bleeding, other adverse events, and liver-function abnormalities | Same as ARISTOTLE away from:<br>- Myocardial infarction, any bleeding, other adverse events, and liver-function abnormalities will not be assessed | Same as target trial<br>Outcomes recorded using ICD-10 codes |
| **Follow-up period** | Start at randomization and ends at the earliest of death, withdrawal of consent to be followed-up, loss to follow-up or the study end date. | Same as ARISTOTLE | Same as target apart from withdrawal of consent |
| **Causal contrasts of interest** | Intention-to-treat for efficacy and safety population for safety | A treatment policy estimand for effectivness and a hypotheical estimand for major bleeding | Observational analogue of specifed estimands |

*(continued)*

| Protocol component | ARISTOTLE | Target trial | Observational analysis |
| --- | --- | --- | --- |
| **Statistical analysis plan** | - Non-inferiority hypothesis required that apixaban upper boundary of the 95% confidence interval for the relative risk would be less than 1.38 for primary efficacy outcome<br>- Relative risk was calculated using Cox proportional-hazards model, with previous warfarin status and geographic region used as strata in the model.<br>- Event rates will be estimated and plotted over time using Kaplan-Meier methodology.<br>- Subgroup analyses:<br>- Prior warfarin/VKA status - Apixaban dose - Geographic Region - Age - Gender - Race - Ethnicity - Weight - Level of Renal Impairment - Number of risk factors - CHADS2 Score - Prior Stroke or TIA - Age $\geq 75$<br>- Diabetes Mellitus - Hypertension requiring pharmacological treatment - Heart Failure - Aspirin at randomization | - Same as ARISTOTLE apart from:<br>- Geographic region will not be used as strata in the model.<br>- The analysis will be stratifed by BMI groups<br>- No subgroup analyses.<br>Risk ratios and risk differences will be estimated through a weighted IPW estimator that coincides with Aalen-Johansen estimator for total effects and a weighted Kaplan-Meier estimator for direct effects<br>Cummulative incidence figures will be reported<br>Non-inferiority margin will not be used due to differences in study population | Same as target trial with adjustment for baseline confounders using inverse probability treatment weighting and for ceonsring inverse probability censoring weighting in each outcome |

### C Variables

**Table S3.** Variables definition in the study

| Variables | Definition | Levels (form used in analysis) | Data source | Outcome ICD-10 Codes |
|---|---|---|---|---|
| Treatments | First prescription of study treatments | 1: Apixaban; 2: Warfarin | CPRD Aurum | |
| Stroke/SE | First event of stroke or systemic embolism during follow-up | 0: No; 1: Yes | HES APC | I60.1, I60.2, I60.3, I60.4, I60.5, I60.6, I60.7, I60.8, I60.9, I61.0, I61.1, I61.2, I61.3, I61.4, I61.5, I61.6, I61.7, I61.8, I61.9, I62.0, I62.1, I62.9, I63.0, I63.1, I63.2, I63.3, I63.4, I63.5, I63.6, I63.8, I63.9, I64, I74.0, I74.1, I74.2, I74.3, I74.4, I74.5, I74.8, I74.9 |

*(continued)*

| Variables | Definition | Levels (form used in analysis) | Data source | Outcome ICD-10 Codes |
|---|---|---|---|---|
| Stroke (any type) | First event of is-chemic/hemorrghic stroke during follow-up | 0: No; 1: Yes | HES APC | I60.1, I60.2, I60.3, I60.4, I60.5, I60.6, I60.7, I60.8, I60.9, I61.0, I61.1, I61.2, I61.3, I61.4, I61.5, I61.6, I61.7, I61.8, I61.9, I62.0, I62.1, I62.9, I63.0, I63.1, I63.2, I63.3, I63.4, I63.5, I63.6, I63.8, I63.9, I64 |
| Ischemic stroke | First event of ischemic stroke during follow-up | 0: No; 1: Yes | HES APC | I63.0, I63.1, I63.2, I63.3, I63.4, I63.5, I63.6, I63.8, I63.9, I64 |
| Hemorrghic stroke | First event of hemorrghic stroke during follow-up | 0: No; 1: Yes | HES APC | I60.1, I60.2, I60.3, I60.4, I60.5, I60.6, I60.7, I60.8, I60.9, I61.0, I61.1, I61.2, I61.3, I61.4, I61.5, I61.6, I61.7, I61.8, I61.9, I62.0, I62.1, I62.9 |

*(continued)*

| Variables | Definition | Levels (form used in analysis) | Data source | Outcome ICD-10 Codes |
|---|---|---|---|---|
| Systematic embolism | First event of systemic embolism during follow-up | 0: No; 1: Yes | HES APC | I74.0, I74.1, I74.2, I74.3, I74.4, I74.5, I74.8, I74.9 |

*(continued)*

| Variables | Definition | Levels (form used in analysis) | Data source | Outcome ICD-10 Codes |
|---|---|---|---|---|
| Major bleeding | First event of major bleeding during follow-up | 0: No; 1: Yes | HES APC | D68.3, H35.6 H43.1, H45.0, I60.0, I60.1, I60.2, I60.3, I60.4, I60.5, I60.6, I60.7, I60.8, I60.9, I61.0, I61.1, I61.2, I61.3, I61.4, I61.5, I61.6, I61.8, I61.9, I62.0, I62.1, I62.9, I85.0, I98.3, K25.0, K25.2, K25.4, K25.6, K26.0, K26.2, K26.4, K26.6, K27.0, K27.4, K27.6, K28.0, K28.2, K28.4, K28.6, K29.0, K62.5, K66.1, K92.0, K92.1, K92.2, M25.0, N02.0, N02.1, N02.2, N02.3, N02.4, N02.5, N02.7, N02.8, N02.9, N93.8, N93.9, N95.0, R04.1, R04.2, R04.8, R04.9, R58 |
| Death | Death within follow-up period | 0: No; 1: Yes | ONS | |

*(continued)*

| Variables | Definition | Levels (form used in analysis) | Data source | Outcome ICD-10 Codes |
|---|---|---|---|---|
| Age | Patient age at enrollment assuming mid year birth | Numeric (Linear, quadratic) | CPRD Aurum | |
| Gender | Patient gender | 1: Male; 2: Female | CPRD Aurum | |
| Individual level index of multiple deprivation (2015) | Individual socioeconmoic status mapped to postcode of residence according to 2015 English Index of Multiple Deprivation | 1 (least deprived), 2, 3, 4, 5 (most deprived) | CPRD Aurum | |
| Practice level index of multiple deprivation (2015) | Practice socioeconmoic status mapped to postcode according to 2015 English Index of Multiple Deprivation | 1 (least deprived), 2, 3, 4, 5 (most deprived) | CPRD Aurum | |

*(continued)*

| Variables | Definition | Levels (form used in analysis) | Data source | Outcome ICD-10 Codes |
|---|---|---|---|---|
| BMI | Lat observation prior to index date | Numeric | CPRD Aurum | |
| BMI groups | Grouped BMI | 1:Normal weight; 2:Overweight; 3:Obese | CPRD Aurum | |
| Smoking status | Any history at baseline | 1:Non-smoker; 2:Ex-smoker; 3:Current-smoker | CPRD Aurum | |
| Alcohol | Alcohol consumption at baseline | 1:Non-drinker; 2:Light-drinker; 3:Moderate-drinker; 4:Heavy-drinker | CPRD Aurum | |
| Ethinic groups | Ethnic group | 1:Black; 2:East Asian; 3:Mixed; 4:Other; 5: South Asian; 6:White | CPRD Aurum | |

*(continued)*

| Variables | Definition | Levels (form used in analysis) | Data source | Outcome ICD-10 Codes |
|---|---|---|---|---|
| Renal function | Renal function based on last creatinine clearance measurement prior to index date | 1:Normal; 2:Mild impairment; 3:Moderate impairment; 4:Severe impairment | CPRD Aurum | |
| Index year | Year of first prescription of study treatments | 1:2013; 2:2014; 3:2015; 4:2016; 5:2017; 6:2018; 7:2019 | CPRD Aurum | |
| Prior stroke | Any history at baseline | 0: No; 1: Yes | CPRD Aurum and HES APC | |
| Prior transient ischemic attack | Any history at baseline | 0: No; 1: Yes | CPRD Aurum and HES APC | |
| Prior systemic embolism | Any history at baseline | 0: No; 1: Yes | CPRD Aurum and HES APC | |

*(continued)*

| Variables | Definition | Levels (form used in analysis) | Data source | Outcome ICD-10 Codes |
|---|---|---|---|---|
| Congestive heart failure or left ventricular dysfunction (LVEF) | Any history at baseline or LVEF value less than or equal to 40% | 0: No; 1: Yes | CPRD Aurum and HES APC | |
| Diabetes | Any history at baseline | 0: No; 1: Yes | CPRD Aurum | |
| Hypertension requrieing treatment | Any history at baseline | 0: No; 1: Yes | CPRD Aurum | |
| History of Prephiral artery disease | Any history at baseline | 0: No; 1: Yes | CPRD Aurum and HES APC | |
| History of Aortic plauge | Any history at baseline | 0: No; 1: Yes | CPRD Aurum and HES APC | |
| History of non-major bleeding | History prior to index date | 0: No; 1: Yes | CPRD Aurum and HES APC | |
| History of myocardial Infarction | Any recored in patient file at baseline | 0: No; 1: Yes | CPRD Aurum and HES APC | |
| History of Chronic obstructive disease | Any recored in patient file at baseline | 0: No; 1: Yes | CPRD Aurum and HES APC | |

*(continued)*

| Variables | Definition | Levels (form used in analysis) | Data source | Outcome ICD-10 Codes |
|---|---|---|---|---|
| History of Liver disease | Any recored in patient file at baseline | 0: No; 1: Yes | CPRD Aurum and HES APC | |
| History of Hematological cancer | Any recored in patient file at baseline | 0: No; 1: Yes | CPRD Aurum and HES APC | |
| History of History of valve surgury | Any recored in patient file at baseline | 0: No; 1: Yes | CPRD Aurum and HES APC | |
| History of Connective tissue diseases | Any recored in patient file at baseline | 0: No; 1: Yes | CPRD Aurum and HES APC | |
| History of Hemiplegia | Any recored in patient file at baseline | 0: No; 1: Yes | CPRD Aurum and HES APC | |
| History of Peptic ulcer | Any recored in patient file at baseline | 0: No; 1: Yes | CPRD Aurum and HES APC | |
| History of solid cancer diagnosis | Any recored in patient file at baseline | 0: No; 1: Yes | CPRD Aurum and HES APC | |
| History of valvular disease | Any recored in patient file at baseline | 0: No; 1: Yes | CPRD Aurum and HES APC | |

*(continued)*

| Variables | Definition | Levels (form used in analysis) | Data source | Outcome ICD-10 Codes |
|---|---|---|---|---|
| Use of Amiodarone | Concomitant use at baseline | 0: No; 1: Yes | CPRD Aurum | |
| Use of aspirin | Concomitant use at baseline | 0: No; 1: Yes | CPRD Aurum | |
| Use of antacids | Concomitant use at baseline | 0: No; 1: Yes | CPRD Aurum | |
| Use of Angiotensin-converting enzyme inhibitors and angiotensin II receptor blockers | Concomitant use at baseline | 0: No; 1: Yes | CPRD Aurum | |
| Use of beta-blocker | Concomitant use at baseline | 0: No; 1: Yes | CPRD Aurum | |
| Use of Clopidogrel | Concomitant use at baseline | 0: No; 1: Yes | CPRD Aurum | |
| Use of digoxin | Concomitant use at baseline | 0: No; 1: Yes | CPRD Aurum | |

*(continued)*

| **Variables** | **Definition** | **Levels (form used in analysis)** | **Data source** | **Outcome ICD-10 Codes** |
|---|---|---|---|---|
| Use of non-steroidal anti-inflammatory drugs | Concomitant use at baseline | 0: No; 1: Yes | CPRD Aurum | |
| Use of Statins | Concomitant use at baseline | 0: No; 1: Yes | CPRD Aurum | |

**BMI:** Body mass Index

### D Directed acyclic graphs

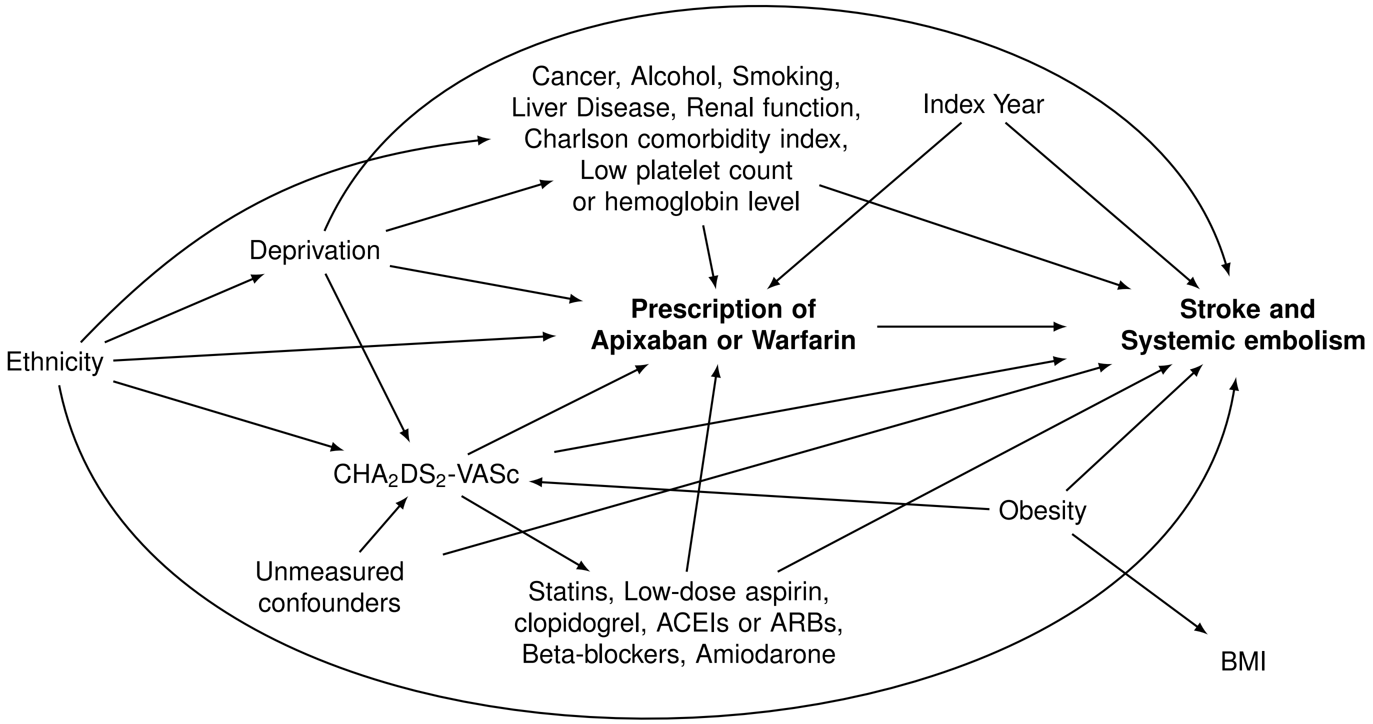


**$CHA_2DS_2$-VASc:** Congestive heart failure, hypertension, age $\geq$ 75, diabetes mellitus, prior stroke, transient ischaemic attack or thromboembolism, vascular disease (peripheral artery disease, myocardial infarction, aortic plaque), age 65 to 74 and sex category (female); **ACEIs:** Angiotensin-converting enzyme inhibitors; **ARBs:** Angiotensin receptor blockers;
**BMI:** Body mass index, an effect modifier by proxy

**Figure S1.** Directed acyclic graph for stroke, systemic embolism outcomes

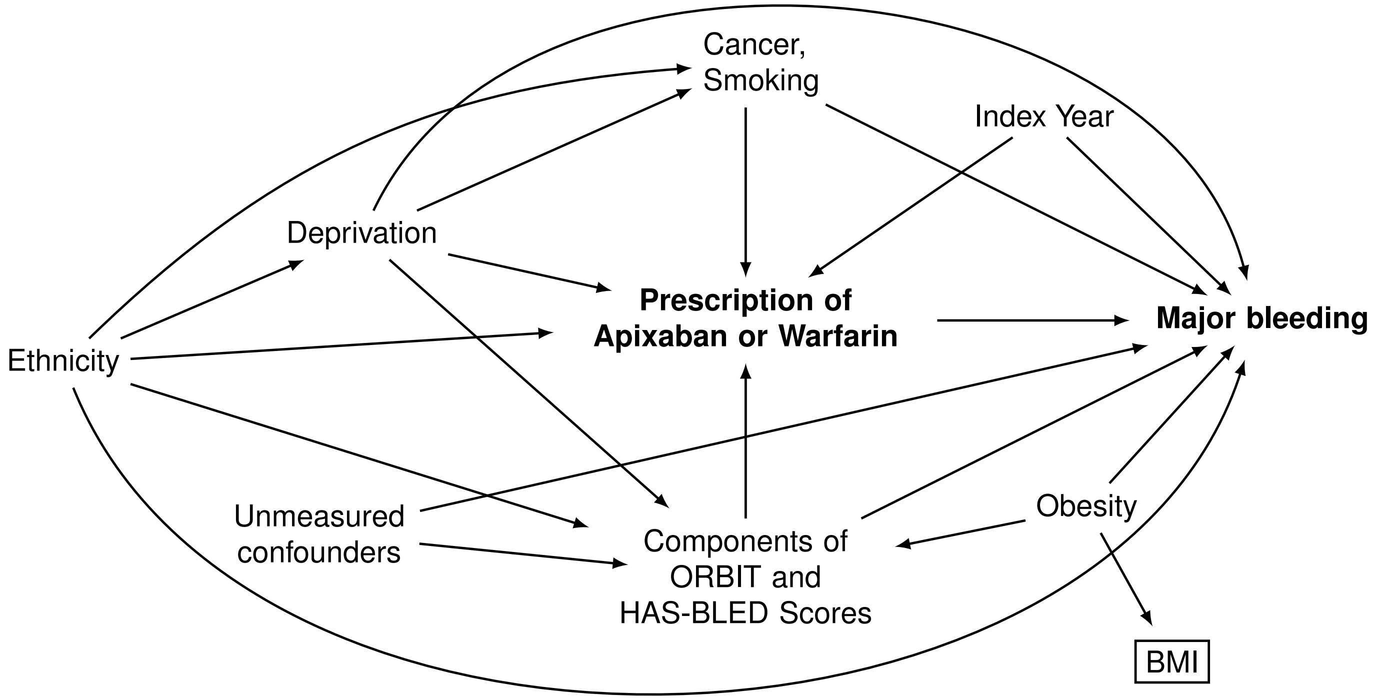


**Components of ORBIT and HAS-BLED Scores:** Low hemoglobin or hematocrit, age $>$ 74 years, any history of GI bleeding, intracranial bleeding, or hemorrhagic stroke, Glomerular filtration rate $<$ 60 mL/min/1.73 $m^2$, using antiplatelet agents (Aspirin, clopidogrel, NSAIDs), hypertension, liver disease and alcohol use;
**BMI:** Body mass index, an effect modifier by proxy

**Figure S2.** Directed acyclic graph for major bleeding

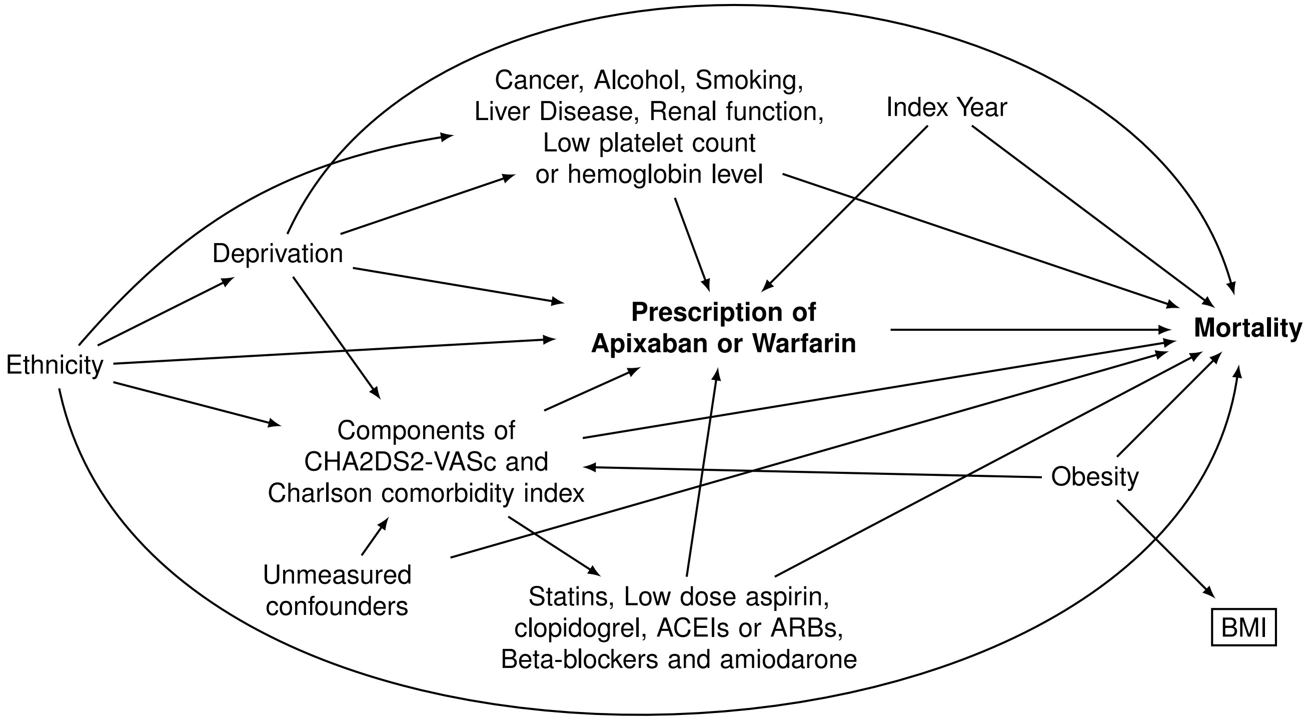


**$CHA_2DS_2$-VASc:** Congestive heart failure (CHF), hypertension, age $\geq$ 75, Diabetes mellitus (DM), prior stroke, transient ischaemic attack (TIA) or thromboembolism, vascular disease (peripheral artery disease (PAD), Myocardial infarction (MI), aortic plaque), age 65 to 74 and sex category (female); **Charlson comorbidity index:** Age, MI, CHF, PAD, prior stroke or TIA, dementia, chronic obstructive pulmonary disease, connective tissue disease, peptic ulcer disease, liver disease, DM, hemiplegia, chronic kidney disease, solid tumor, leukemia and lymphoma; **BMI:** Body mass index, an effect modifier by proxy

**Figure S3.** Directed acyclic graph for mortality

**Table S4.** Baseline characteristics of eligible study participants using CPRD linked data by treatment group with unweighted and inverse probability treatment weighted standardised mean differences

| Characteristics | Apixaban | Warfarin | Overall | Unweighted SMD | IPT weighted SMD |
|---|---|---|---|---|---|
| **Number of patients (N)** | 27747 | 23145 | 50892 | | |
| **Demographics** | | | | | |
| **Age at entry** (years)-median [IQR] | 78.00 [71.00, 84.00] | 77.00 [71.00, 82.00] | 78.00 [71.00, 83.00] | 0.149 | 0.042 |
| **Age groups at entry** (years)-N (%) | | | | 0.072 | 0.097 |
| 18-49 | 118 (0.4) | 116 (0.5) | 234 (0.5) | | |
| 50-59 | 744 (2.7) | 673 (2.9) | 1417 (2.8) | | |
| 60-69 | 3545 (12.8) | 3383 (14.6) | 6928 (13.6) | | |
| 70-74 | 5397 (19.5) | 4776 (20.6) | 10173 (20.0) | | |
| $\geq$ 75 | 17943 (64.7) | 14197 (61.3) | 32140 (63.2) | | |
| **Female**- N (%) | 13079 (47.1) | 10363 (44.8) | 23442 (46.1) | 0.047 | 0.008 |
| **Weight** (kg)- median [IQR] | 79.00 [67.00, 92.00] | 81.00 [69.60, 94.00] | 80.00 [68.00, 93.00] | 0.111 | 0.132 |
| **BMI** (kg/m$^2$)- median [IQR] | 27.80 [24.60, 31.80] | 28.30 [25.00, 32.32] | 28.00 [24.80, 32.00] | 0.087 | 0.133 |
| **BMI groups** (kg/m$^2$)- N (%) | | | | 0.088 | 0.120 |

*(continued)*

| Characteristics | Apixaban | Warfarin | Overall | Unweighted SMD | IPT weighted SMD |
|---|---|---|---|---|---|
| Underweight <18.5 | 438 (1.6) | 245 (1.1) | 683 (1.3) | | |
| Normal weight 18.5-24.9 | 7231 (26.1) | 5446 (23.5) | 12677 (24.9) | | |
| Overweight 25-29.9 | 10280 (37.0) | 8561 (37.0) | 18841 (37.0) | | |
| Obesity class I 30-34.9 | 6065 (21.9) | 5368 (23.2) | 11433 (22.5) | | |
| Obesity class II 35-39.9 | 2396 (8.6) | 2228 (9.6) | 4624 (9.1) | | |
| Obesity class III ≥ 40 | 1337 (4.8) | 1297 (5.6) | 2634 (5.2) | | |
| Individual-level IMD(2015)- N (%) | | | | 0.044 | 0.029 |
| 1st quintile (Least deprived) | 6924 (25.0) | 5529 (23.9) | 12453 (24.5) | | |
| 2nd quintile | 6446 (23.2) | 5346 (23.1) | 11792 (23.2) | | |
| 3rd quintile | 5401 (19.5) | 4692 (20.3) | 10093 (19.8) | | |
| 4th quintile | 4638 (16.7) | 4132 (17.9) | 8770 (17.2) | | |
| 5th quintile (Most deprived) | 4338 (15.6) | 3446 (14.9) | 7784 (15.3) | | |
| Practice-level IMD(2015)- N (%) | | | | 0.054 | 0.021 |
| 1st quintile (Least deprived) | 5342 (19.3) | 4285 (18.5) | 9627 (18.9) | | |

*(continued)*

| **Characteristics** | **Apixaban** | **Warfarin** | **Overall** | **Unweighted SMD** | **IPT weighted SMD** |
|---|---|---|---|---|---|
| 2nd quintile | 5704 (20.6) | 4573 (19.8) | 10277 (20.2) | | |
| 3rd quintile | 5205 (18.8) | 4822 (20.8) | 10027 (19.7) | | |
| 4th quintile | 6021 (21.7) | 4922 (21.3) | 10943 (21.5) | | |
| 5th quintile (Most deprived) | 5475 (19.7) | 4543 (19.6) | 10018 (19.7) | | |
| Ethnicity- N (%) | | | | 0.023 | 0.022 |
| Black | 276 (1.0) | 283 (1.2) | 559 (1.1) | | |
| East Asian | 37 (0.1) | 31 (0.1) | 68 (0.1) | | |
| Mixed | 62 (0.2) | 52 (0.2) | 114 (0.2) | | |
| Other | 71 (0.3) | 53 (0.2) | 124 (0.2) | | |
| South Asian | 524 (1.9) | 445 (1.9) | 969 (1.9) | | |
| White | 26777 (96.5) | 22281 (96.3) | 49058 (96.4) | | |
| **Lifestyle factors**- N (%) | | | | | |
| Smoking status | | | | 0.055 | 0.008 |
| Non-smoker | 10190 (36.7) | 7942 (34.3) | 18132 (35.6) | | |
| Former smoker | 15550 (56.0) | 13589 (58.7) | 29139 (57.3) | | |
| Current smoker | 2007 (7.2) | 1614 (7.0) | 3621 (7.1) | | |

*(continued)*

| Characteristics | Apixaban | Warfarin | Overall | Unweighted SMD | IPT weighted SMD |
|---|---|---|---|---|---|
| Alcohol consumption | | | | 0.060 | 0.017 |
| Non-drinker | 10631 (38.3) | 8453 (36.5) | 19084 (37.5) | | |
| Light drinker | 12972 (46.8) | 11425 (49.4) | 24397 (47.9) | | |
| Moderate | 3537 (12.7) | 2872 (12.4) | 6409 (12.6) | | |
| Heavy | 607 (2.2) | 395 (1.7) | 1002 (2.0) | | |
| **Disease charactirsitcis-** N (%) | | | | | |
| $CHA_2DS_2$-VASc components | | | | | |
| Congestive heart failure (or left ventricular systolic dysfunction) | 6529 (23.5) | 5164 (22.3) | 11693 (23.0) | 0.029 | 0.018 |
| Treated hypertension | 21451 (77.3) | 17969 (77.6) | 39420 (77.5) | 0.008 | 0.002 |
| Systolic blood pressure (mm Hg)- median [IQR] | 132.00 [121.00, 142.00] | 132.00 [121.00, 140.00] | 132.00 [121.00, 141.00] | 0.016 | 0.002 |
| Diabetes mellitus | 8051 (29.0) | 6663 (28.8) | 14714 (28.9) | 0.005 | 0.019 |
| Stroke, TIA, or SE | 7356 (26.5) | 4928 (21.3) | 12284 (24.1) | 0.123 | 0.027 |
| **History of vascular diseases** | | | | | |

*(continued)*

| **Characteristics** | **Apixaban** | **Warfarin** | **Overall** | **Unweighted SMD** | **IPT weighted SMD** |
|---|---|---|---|---|---|
| Peripheral artery disease | 1803 (6.5) | 1600 (6.9) | 3403 (6.7) | 0.017 | 0.019 |
| Aortic Plaque | 5817 (21.0) | 5053 (21.8) | 10870 (21.4) | 0.021 | 0.016 |
| Myocardial infarction | 3582 (12.9) | 3031 (13.1) | 6613 (13.0) | 0.006 | 0.010 |
| $CHA_2DS_2$-VASc score | | | | 0.114 | 0.051 |
| 2 | 5956 (21.5) | 5385 (23.3) | 11341 (22.3) | | |
| 3 | 8221 (29.6) | 7446 (32.2) | 15667 (30.8) | | |
| 4 | 5750 (20.7) | 4883 (21.1) | 10633 (20.9) | | |
| 5 | 4342 (15.6) | 3163 (13.7) | 7505 (14.7) | | |
| 6 | 2297 (8.3) | 1523 (6.6) | 3820 (7.5) | | |
| 7 | 921 (3.3) | 586 (2.5) | 1507 (3.0) | | |
| 8 | 260 (0.9) | 159 (0.7) | 419 (0.8) | | |
| **Non-major bleeding** | 7806 (28.1) | 5826 (25.2) | 13632 (26.8) | 0.067 | 0.016 |
| **Fall in previous year** | 789 (2.8) | 287 (1.2) | 1076 (2.1) | 0.114 | 0.041 |
| **Comorbidities**- N (%) | | | | | |
| Renal function | | | | 0.108 | 0.056 |
| Normal | 7946 (28.6) | 7607 (32.9) | 15553 (30.6) | | |

*(continued)*

| Characteristics | Apixaban | Warfarin | Overall | Unweighted SMD | IPT weighted SMD |
|---|---|---|---|---|---|
| Mild impairment | 12093 (43.6) | 9841 (42.5) | 21934 (43.1) | | |
| Moderate impairment | 6307 (22.7) | 4480 (19.4) | 10787 (21.2) | | |
| Severe impairment | 1401 (5.0) | 1217 (5.3) | 2618 (5.1) | | |
| Charlson comorbidity index components | | | | | |
| COPD | 3820 (13.8) | 2896 (12.5) | 6716 (13.2) | 0.037 | 0.030 |
| Connective tissue disease | 1933 (7.0) | 1471 (6.4) | 3404 (6.7) | 0.025 | 0.016 |
| Peptic ulcer disease | 1485 (5.4) | 1263 (5.5) | 2748 (5.4) | 0.005 | 0.024 |
| Liver disease | 236 (0.9) | 161 (0.7) | 397 (0.8) | 0.018 | 0.001 |
| Hemiplegia | 86 (0.3) | 42 (0.2) | 128 (0.3) | 0.026 | 0.016 |
| Non-hematological cancer | 4385 (15.8) | 3447 (14.9) | 7832 (15.4) | 0.025 | 0.009 |
| Hematological cancer | 602 (2.2) | 481 (2.1) | 1083 (2.1) | 0.006 | 0.006 |
| Concomitant medications at index date | | | | | |
| ACE inhibitors or ARBs | 14083 (50.8) | 12838 (55.5) | 26921 (52.9) | 0.095 | 0.011 |
| Amiodarone | 482 (1.7) | 485 (2.1) | 967 (1.9) | 0.026 | 0.008 |
| Antacids | 719 (2.6) | 598 (2.6) | 1317 (2.6) | 0.001 | 0.002 |
| Aspirin | 2274 (8.2) | 3282 (14.2) | 5556 (10.9) | 0.191 | 0.036 |
| Beta-blockers | 17429 (62.8) | 14071 (60.8) | 31500 (61.9) | 0.042 | 0.009 |

*(continued)*

| Characteristics | Apixaban | Warfarin | Overall | Unweighted SMD | IPT weighted SMD |
|---|---|---|---|---|---|
| Calcium channel blockers | 8800 (31.7) | 7808 (33.7) | 16608 (32.6) | 0.043 | 0.044 |
| Clopidogrel | 996 (3.6) | 1087 (4.7) | 2083 (4.1) | 0.056 | 0.010 |
| Digoxin | 2562 (9.2) | 2345 (10.1) | 4907 (9.6) | 0.030 | 0.074 |
| NSAIDs | 2096 (7.6) | 1487 (6.4) | 3583 (7.0) | 0.044 | 0.025 |
| Statins | 15867 (57.2) | 13305 (57.5) | 29172 (57.3) | 0.006 | 0.008 |
| Index year- N (%) | | | | 1.893 | 0.035 |
| 2013 | 170 (0.6) | 7150 (30.9) | 7320 (14.4) | | |
| 2014 | 1030 (3.7) | 6297 (27.2) | 7327 (14.4) | | |
| 2015 | 2920 (10.5) | 4697 (20.3) | 7617 (15.0) | | |
| 2016 | 4973 (17.9) | 2672 (11.5) | 7645 (15.0) | | |
| 2017 | 6623 (23.9) | 1327 (5.7) | 7950 (15.6) | | |
| 2018 | 7387 (26.6) | 676 (2.9) | 8063 (15.8) | | |
| 2019 | 4644 (16.7) | 326 (1.4) | 4970 (9.8) | | |

**ACE inhibitors or ARBs:** Angiotensin-converting enzyme inhibitors and angiotensin II receptor blockers; **BMI:** Body mass Index; **$CHA_2DS_2$-VASc score:** calculated as 1 for each component expect for age $\geq$ 75 and history of stroke counted as 2; **COPD:** Chronic obstructive pulmonary disease;**IMD(2015):** Index of Multiple Deprivation 2015; **IQR:** Interquartile range; **IPT weighted:** Inverse probability treatment weighted ; **NSAIDs:** Non-steroidal anti-inflammatory drugs; **SMD:** Standardised mean differences; **SE:** Systemic embolism; **TIA:** Transient ischemic attack

### E Quantitative variables

The quantitative variables were age, systolic blood pressure, weight, and BMI.

Age (years) was reported as continuous variable and was also categorized into 18-49, 50-59, 60-69, 70-74, $\leq$ 75. Unequal intervals were used since the disease is uncommon in younger ages and being more than 75 years constitutes a major reason for initiating anticoagulant treatment.

Systolic blood pressure, and weight were reported as continuous variables.

BMI (kg/m$^2$) was reported as continuous variable and categorized based on NICE's classification[79]:

- Underweight: less than 18.5
- Healthy weight: 18.5-24.9
- Overweight: 25-29.9
- Obesity class I: 30-34.9
- Obesity class II: 35-39.9
- Obesity class III: $\geq$ 40

However, based on the initial estimates of total participants in each BMI group across treatment groups, we regrouped the variable in the analysis to the following categories to avoid data sparsity:

- Healthy weight: $\leq$ 24.9
- Overweight: 25-29.9
- Obese: $\geq$ 30

#### F Inverse probability weights

We first identified the stabilized inverse probability treatment weights, $\mathrm{SW}^{\mathrm{A}}$, for each individual in our study population. The denominator is the probability that an individual received the observed treatment (A), given observed confounders (L). We used different numerators for overall and BMI-stratified results. For overall results, the numerator is the probability of receiving the observed treatment (A). For stratified results, the numerator is the probability of receiving the observed treatment (A) given the effect modifier (V).

- Overall analysis:

$$SW^A(V) = \frac{f(A)}{f(A|L,V)}$$

- Stratified analysis:

$$SW^A(V) = \frac{f(A|V)}{f(A|L,V)}$$

We then identified the stabilized inverse probability censoring weights, $\mathrm{SW}^{\mathrm{C}}$, for each individual in our study population. The denominator is the probability that an individual is not censored (C) given the observed treatment (A) and observed confounders (L). The numerator is the probability that the individual is not censored (C) given the observed treatment (A).

$$SW^C = \frac{f(C=0|A)}{f(C=0|A,L)}$$

We multiplied the weights from the two models to get the final weights $\mathrm{SW}^{\mathrm{A,C}}$. These were used to non-parametrically estimate the cause-specific cumulative incidence described in Young et. al.[20]

$$SW^{A,C} = SW^A(V)XSW^C$$

**Table S5.** Summary of the inverse probability weights for the composite effectiveness outcome of stroke/systemic embolism and major bleeding

| Inverse probability weights (IPW) | Apixaban | | | Warfarin | | |
|---|---|---|---|---|---|---|
| | **Min** | **Max** | **Mean** | **Min** | **Max** | **Mean** |
| **Composite of stroke/systemic embolism** | | | | | | |
| Stabilized Inverse probability treatment weights | 0.53 | 65.98 | 1.21 | 0.43 | 22.87 | 0.82 |
| Stabilized Inverse probability treatment weights (truncated to the 99th percentile) | 0.53 | 6.72 | 1.07 | 0.43 | 6.72 | 0.81 |
| Stabilized Inverse probability censoring weights | 0.30 | 9.64 | 0.94 | 0.86 | 7.29 | 0.99 |
| Stabilized Inverse probability censoring weight (truncated to the 99th percentile) | 0.34 | 2.14 | 0.92 | 0.86 | 2.14 | 0.99 |
| **Major bleeding** | | | | | | |
| Stabilized Inverse probability treatment weights | 0.53 | 64.9 | 1.20 | 0.43 | 23.44 | 0.82 |
| Stabilized Inverse probability treatment weights (truncated to the 99th percentile) | 0.53 | 6.74 | 1.06 | 0.43 | 6.74 | 0.81 |
| Stabilized Inverse probability censoring weights | 0.26 | 11.34 | 0.94 | 0.86 | 4.76 | 0.99 |
| Stabilized Inverse probability censoring weight (truncated to the 99th percentile) | 0.32 | 1.97 | 0.92 | 0.86 | 1.97 | 0.92 |

### G Results of secondary effectiveness outcomes

**Stroke (any type) (IPW)***

| BMI groups | Apixaban (No.= 27,747) Events/No. | Risk** (95 %CI) | RR (95 %CI) | Warfarin (No.= 23,145) Events/No. | Risk** (95 %CI) | RR (95 %CI) | 36-month Risk Ratio (95% CI) |
|---|---|---|---|---|---|---|---|
| Overall | 651/27,747 | 3.8 (2.9,4.7) | | 715/23,145 | 3.2 (2.4,4.1) | | 1.17 (0.82,1.66) |
| Normal weight ≤24.9 kg/m2 | 209/7,669 | 4.2 (3.2,5.2) | 1.19 (0.82,1.71) | 211/5,691 | 3.5 (2.7,4.4) | Reference | 1.19 (0.82,1.71) |
| Overweight 25-29.9 kg/m2 | 271/10,280 | 3.8 (3.1,4.4) | 1.07 (0.77,1.48) | 263/8,561 | 3.5 (2.0,5.0) | 0.99 (0.60,1.63) | 1.08 (0.69,1.70) |
| Obese ≥30 kg/m2 | 171/9,798 | 3.4 (1.4,5.4) | 0.97 (0.56,1.66) | 241/8,893 | 2.7 (2.0,3.5) | 0.77 (0.51,1.17) | 1.25 (0.71,2.22) |

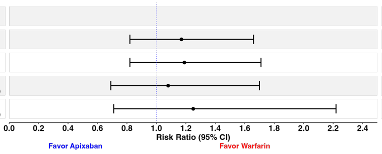


**Ischemic Stroke (IPW)***

| BMI groups | Apixaban (No.= 27,747) Events/No. | Risk** (95 %CI) | RR (95 %CI) | Warfarin (No.= 23,145) Events/No. | Risk** (95 %CI) | RR (95 %CI) | 36-month Risk Ratio (95% CI) |
|---|---|---|---|---|---|---|---|
| Overall | 518/27,747 | 3.0 (1.2,4.8) | | 499/23,145 | 2.2 (1.5,2.9) | | 1.39 (0.90,2.16) |
| Normal weight ≤24.9 kg/m2 | 159/7,669 | 2.9 (2.3,3.5) | 1.22 (0.76,1.95) | 147/5,691 | 2.4 (1.4,3.3) | Reference | 1.22 (0.76,1.95) |
| Overweight 25-29.9 kg/m2 | 225/10,280 | 3.2 (2.7,3.8) | 1.37 (0.89,2.10) | 181/8,561 | 2.4 (0.1,4.9) | 1.02 (0.52,2.01) | 1.34 (0.72,2.47) |
| Obese ≥30 kg/m2 | 134/9,798 | 2.9 (0.9,4.9) | 1.22 (0.65,2.29) | 171/8,893 | 1.8 (0.9,2.6) | 0.75 (0.39,1.41) | 1.64 (0.77,3.48) |

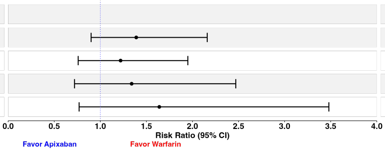


**Haemorrhagic Stroke (IPW)***

| BMI groups | Apixaban (No.= 27,747) Events/No. | Risk** (95 %CI) | RR (95 %CI) | Warfarin (No.= 23,145) Events/No. | Risk** (95 %CI) | RR (95 %CI) | 36-month Risk Ratio (95% CI) |
|---|---|---|---|---|---|---|---|
| Overall | 141/27,747 | 0.8 (0.5,1.0) | | 231/23,145 | 1.1 (0.8,1.4) | | 0.69 (0.46,1.04) |
| Normal weight ≤24.9 kg/m2 | 51/7,669 | 1.3 (0.4,2.3) | 1.09 (0.54,2.21) | 68/5,691 | 1.2 (0.8,1.6) | Reference | 1.09 (0.54,2.21) |
| Overweight 25-29.9 kg/m2 | 51/10,280 | 0.6 (0.4,0.8) | 0.48 (0.28,0.82) | 89/8,561 | 1.2 (0.7,1.6) | 0.95 (0.52,1.74) | 0.50 (0.29,0.86) |
| Obese ≥30 kg/m2 | 39/9,798 | 0.6 (0.4,0.8) | 0.45 (0.25,0.80) | 74/8,893 | 1.0 (0.6,1.5) | 0.82 (0.42,1.60) | 0.55 (0.28,1.08) |

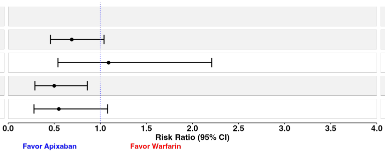


**Systemic embolism (IPW)***

| BMI groups | Apixaban (No.= 27,747) Events/No. | Risk** (95 %CI) | RR (95 %CI) | Warfarin (No.= 23,145) Events/No. | Risk** (95 %CI) | RR (95 %CI) | 36-month Risk Ratio (95% CI) |
|---|---|---|---|---|---|---|---|
| Overall | 68/27,747 | 0.5 (0.3,0.7) | | 130/23,145 | 0.5 (0.3,0.7) | | 0.95 (0.51,1.74) |
| Normal weight ≤24.9 kg/m2 | 27/7,669 | 0.5 (0.3,0.7) | 0.90 (0.39,2.06) | 32/5,691 | 0.6 (0.2,0.9) | Reference | 0.90 (0.39,2.06) |
| Overweight 25-29.9 kg/m2 | 21/10,280 | 0.5 (0.2,0.8) | 0.89 (0.33,2.41) | 56/8,561 | 0.5 (0.3,0.7) | 0.96 (0.42,2.18) | 0.93 (0.36,2.36) |
| Obese ≥30 kg/m2 | 20/9,798 | 0.5 (0.1,0.9) | 0.91 (0.27,3.10) | 42/8,893 | 0.5 (0.2,0.9) | 0.92 (0.35,2.38) | 1.00 (0.28,3.54) |

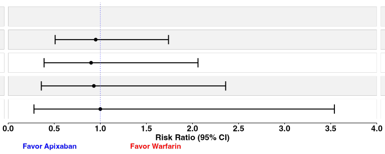


**Stroke (any type) (IPW)***

| BMI groups | Events/No. | Risk** (95 %CI) | RD (95 %CI) | Events/No. | Risk** (95 %CI) | RD (95 %CI) | 36-month Risk difference (95% CI) |
|---|---|---|---|---|---|---|---|
| Overall | 651/27,747 | 3.8 (2.9,4.7) | | 715/23,145 | 3.2 (2.4,4.1) | | 0.5 (-0.7,1.8) |
| Normal weight ≤24.9 kg/m2 | 209/7,669 | 4.2 (3.2,5.2) | 0.7 (-0.6,1.9) | 211/5,691 | 3.5 (2.7,4.4) | Reference | 0.7 (-0.6,1.9) |
| Overweight 25-29.9 kg/m2 | 271/10,280 | 3.8 (3.1,4.4) | 0.2 (-0.8,1.3) | 263/8,561 | 3.5 (2.0,5.0) | 0.0 (-1.7,1.7) | 0.3 (-1.3,1.9) |
| Obese ≥30 kg/m2 | 171/9,798 | 3.4 (1.4,5.4) | -0.1 (-2.3,2.0) | 241/8,893 | 2.7 (2.0,3.5) | -0.8 (-1.9,0.3) | 0.7 (-1.5,2.9) |

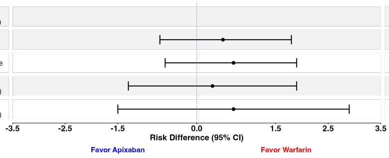


**Ischemic Stroke (IPW)***

| BMI groups | Events/No. | Risk** (95 %CI) | RD (95 %CI) | Events/No. | Risk** (95 %CI) | RD (95 %CI) | 36-month Risk difference (95% CI) |
|---|---|---|---|---|---|---|---|
| Overall | 518/27,747 | 3.0 (1.2,4.8) | | 499/23,145 | 2.2 (1.5,2.9) | | 0.9 (-1.1,2.8) |
| Normal weight ≤24.9 kg/m2 | 159/7,669 | 2.9 (2.3,3.5) | 0.5 (-0.6,1.7) | 147/5,691 | 2.4 (1.4,3.3) | Reference | 0.5 (-0.6,1.7) |
| Overweight 25-29.9 kg/m2 | 225/10,280 | 3.2 (2.7,3.8) | 0.9 (-0.2,1.9) | 181/8,561 | 2.4 (0.1,4.9) | 0.1 (-2.6,2.7) | 0.8 (-1.8,3.5) |
| Obese ≥30 kg/m2 | 134/9,798 | 2.9 (0.9,4.9) | 0.5 (-1.6,2.6) | 171/8,893 | 1.8 (0.9,2.6) | -0.6 (-1.9,0.7) | 1.1 (-1.1,3.4) |

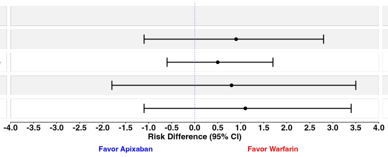


**Haemorrhagic Stroke (IPW)***

| BMI groups | Events/No. | Risk** (95 %CI) | RD (95 %CI) | Events/No. | Risk** (95 %CI) | RD (95 %CI) | 36-month Risk difference (95% CI) |
|---|---|---|---|---|---|---|---|
| Overall | 141/27,747 | 0.8 (0.5,1.0) | | 231/23,145 | 1.1 (0.8,1.4) | | -0.3 (-0.7,0.0) |
| Normal weight ≤24.9 kg/m2 | 51/7,669 | 1.3 (0.4,2.3) | 0.1 (-1.0,1.2) | 68/5,691 | 1.2 (0.8,1.6) | Reference | 0.1 (-1.0,1.2) |
| Overweight 25-29.9 kg/m2 | 51/10,280 | 0.6 (0.4,0.8) | -0.6 (-1.1,-0.2) | 89/8,561 | 1.2 (0.7,1.6) | -0.1 (-0.7,0.6) | -0.6 (-1.1,-0.1) |
| Obese ≥30 kg/m2 | 39/9,798 | 0.6 (0.4,0.8) | -0.7 (-1.1,-0.2) | 74/8,893 | 1.0 (0.6,1.5) | -0.2 (-0.8,0.4) | -0.5 (-0.9,0.0) |

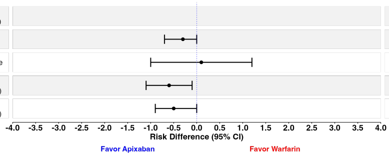


**Systemic embolism (IPW)***

| BMI groups | Events/No. | Risk** (95 %CI) | RD (95 %CI) | Events/No. | Risk** (95 %CI) | RD (95 %CI) | 36-month Risk difference (95% CI) |
|---|---|---|---|---|---|---|---|
| Overall | 68/27,747 | 0.5 (0.3,0.7) | | 130/23,145 | 0.5 (0.3,0.7) | | 0.0 (-0.3,0.3) |
| Normal weight ≤24.9 kg/m2 | 27/7,669 | 0.5 (0.3,0.7) | -0.1 (-0.5,0.4) | 32/5,691 | 0.6 (0.2,0.9) | Reference | -0.1 (-0.5,0.4) |
| Overweight 25-29.9 kg/m2 | 21/10,280 | 0.5 (0.2,0.8) | -0.1 (-0.5,0.4) | 56/8,561 | 0.5 (0.3,0.7) | 0.0 (-0.5,0.4) | 0.0 (-0.4,0.4) |
| Obese ≥30 kg/m2 | 20/9,798 | 0.5 (0.1,0.9) | 0.0 (-0.6,0.5) | 42/8,893 | 0.5 (0.2,0.9) | 0.0 (-0.5,0.4) | 0.0 (-0.5,0.5) |

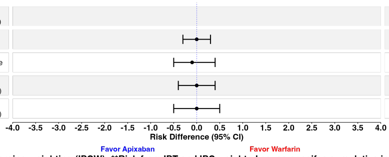


***Using stabilized weights from inverse probability treatment weighting (IPTW) and inverse probablity censroing weighting (IPCW); **Risk from IPT and IPC weighted cause-specifec cumulative incidence estimator;**
**BMI:Body mass index; IPW: Inverse probability weighting; No.: Number of individuals in the analysis; RD:Risk difference; RR:Risk ratio; SE: Systemic Embolism;**
**95% CI: 95% confidence interval estimated from non-parametric bootstrapping using 500 samples**

**Figure S4.** Inverse probability weighted estimates of the 3-year BMI stratum-specific risks, risk differences per 100 people, and risk ratios of the comparative effectiveness for the total effect of apixaban versus warfarin in stroke (any type), ischemic stroke, haemorrhagic stroke, and systemic embolism using a complete-case analysis (Estimands 3,4,5,6)

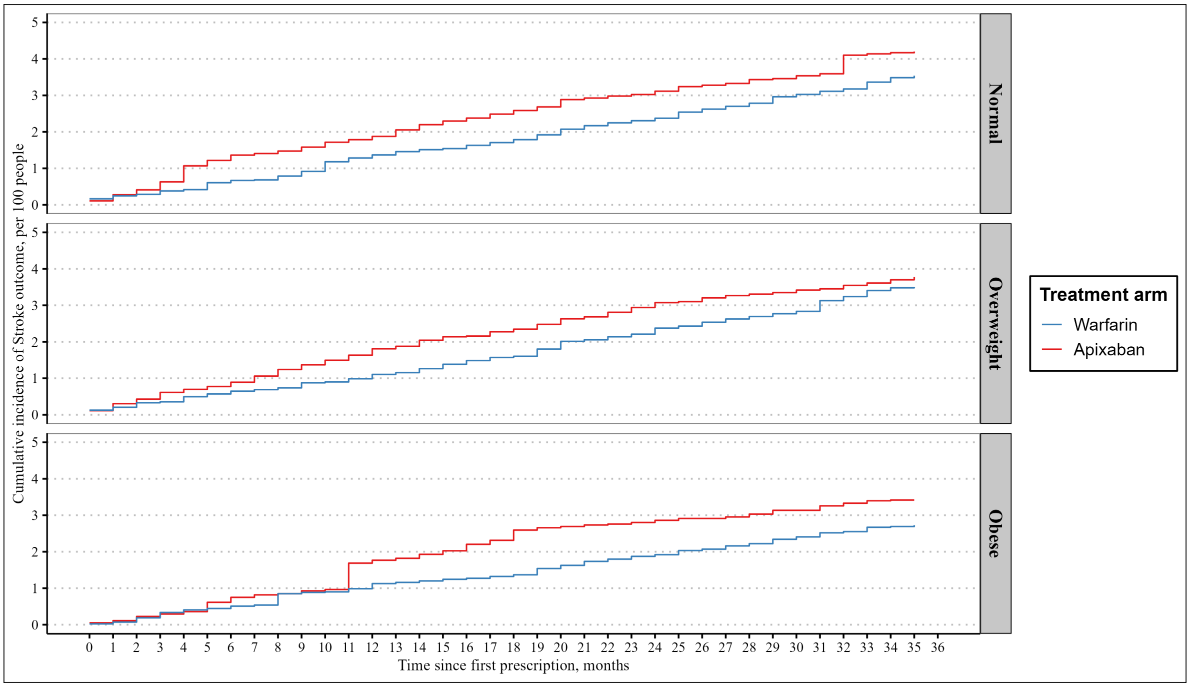


**Figure S5.** Inverse probability weighted cumulative incidence of the 3-year risk of the total effect of apixaban versus warfarin in stroke (ischemic or haemorrhagic) stratified by BMI using a complete-case analysis (Estimand 3)

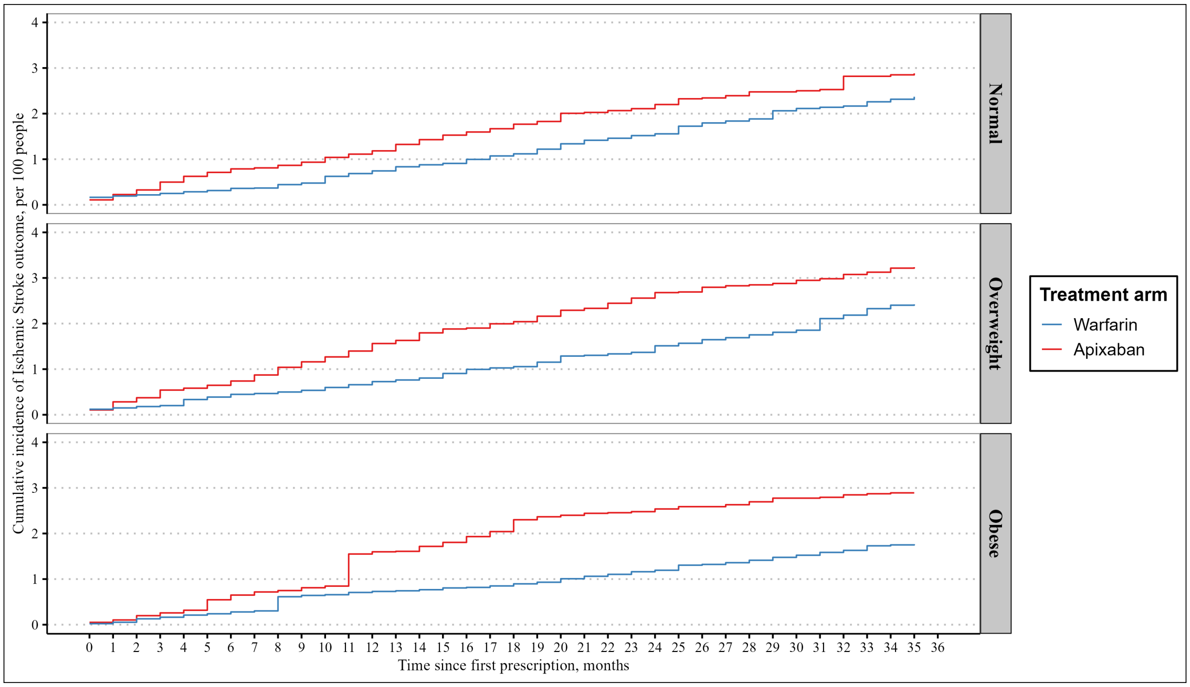


**Figure S6.** Inverse probability weighted cumulative incidence of the 3-year risk of the total effect of apixaban versus warfarin in ischemic stroke stratified by BMI using a complete-case analysis (Estimand 4)

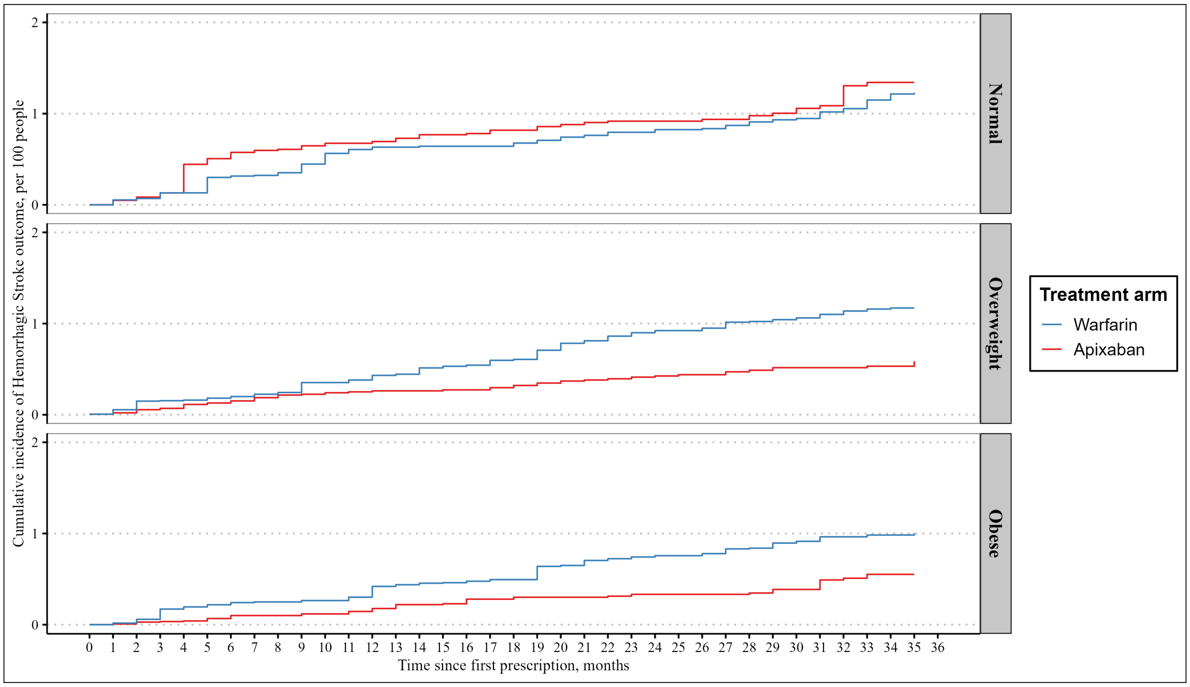


**Figure S7.** Inverse probability weighted cumulative incidence of the 3-year risk of the total effect of apixaban versus warfarin in haemorrhagic stroke stratified by BMI using a complete-case analysis (Estimands 5)

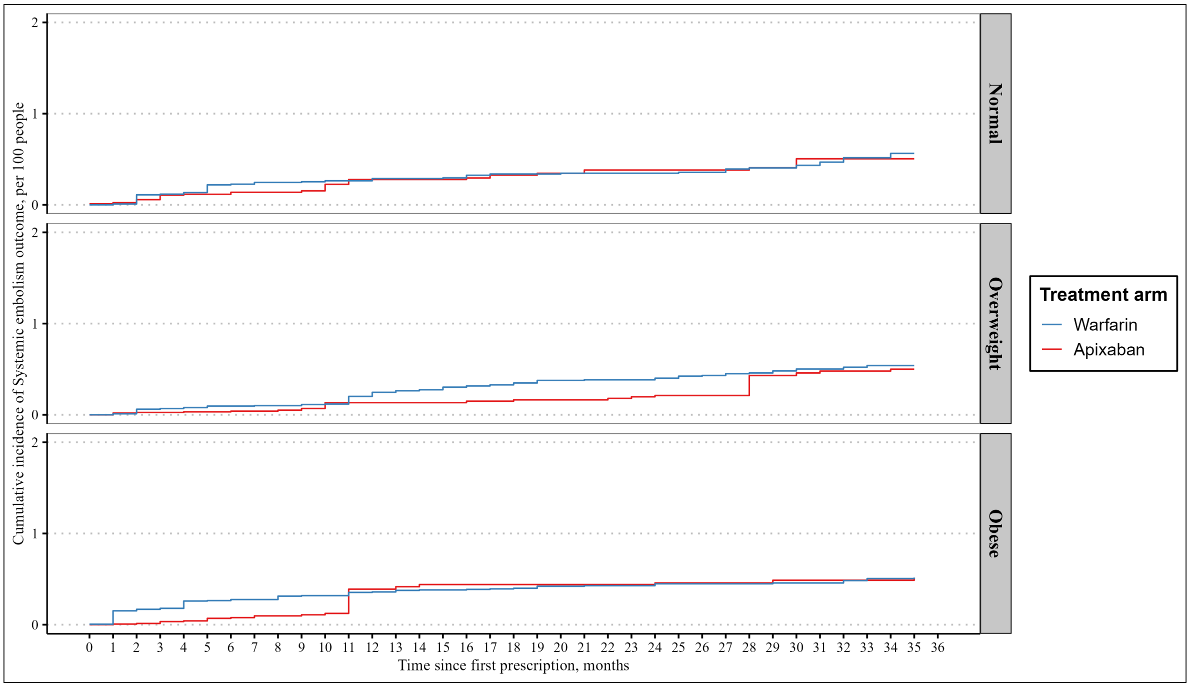


**Figure S8.** Inverse probability weighted cumulative incidence of the 3-year risk of the total effect of apixaban versus warfarin in systemic embolism stratified by BMI using a complete-case analysis (Estimands 6)

### H Supplementary and sensitivity analyses

#### H.1 Supplementary analysis

Stroke/SE (IPW)*

| BMI groups | Apixaban (No.= 27,747) Events/No. | Risk** (95 %CI) | RR (95 %CI) | Warfarin (No.= 23,145) Events/No. | Risk** (95 %CI) | RR (95 %CI) | 36-month Risk Ratio (95% CI) |
|---|---|---|---|---|---|---|---|
| Overall | 719/27,747 | 3.8 (2.6,5.1) | | 842/23,145 | 3.8 (3.1,4.5) | | 1.01 (0.75,1.36) |
| Normal weight ≤24.9 kg/m2 | 236/7,669 | 4.6 (3.7,5.6) | 1.10 (0.80,1.53) | 243/5,691 | 4.2 (3.3,5.1) | Reference | 1.10 (0.80,1.53) |
| Overweight 25-29.9 kg/m2 | 292/10,280 | 3.9 (3.1,4.7) | 0.94 (0.68,1.28) | 318/8,561 | 4.1 (2.9,5.2) | 0.97 (0.67,1.40) | 0.97 (0.69,1.36) |
| Obese ≥30 kg/m2 | 191/9,798 | 3.1 (1.5,4.7) | 0.75 (0.48,1.16) | 281/8,893 | 3.2 (2.3,4.1) | 0.77 (0.53,1.14) | 0.97 (0.59,1.58) |

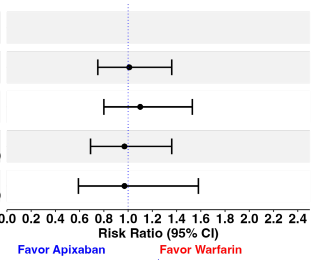


Stroke/SE (IPW)*

| BMI groups | Events/No. | Risk** (95 %CI) | RD (95 %CI) | Events/No. | Risk** (95 %CI) | RD (95 %CI) | 36-month Risk difference (95% CI) |
|---|---|---|---|---|---|---|---|
| Overall | 719/27,747 | 3.8 (2.6,5.1) | | 842/23,145 | 3.8 (3.1,4.5) | | 0.0 (-1.4,1.5) |
| Normal weight ≤24.9 kg/m2 | 236/7,669 | 4.6 (3.7,5.6) | 0.4 (-0.9,1.7) | 243/5,691 | 4.2 (3.3,5.1) | Reference | 0.4 (-0.9,1.7) |
| Overweight 25-29.9 kg/m2 | 292/10,280 | 3.9 (3.1,4.7) | -0.3 (-1.5,0.9) | 318/8,561 | 4.1 (2.9,5.2) | -0.1 (-1.6,1.3) | -0.1 (-1.5,1.2) |
| Obese ≥30 kg/m2 | 191/9,798 | 3.1 (1.5,4.7) | -1.1 (-2.8,0.7) | 281/8,893 | 3.2 (2.3,4.1) | -1.0 (-2.3,0.4) | -0.1 (-2.0,1.8) |

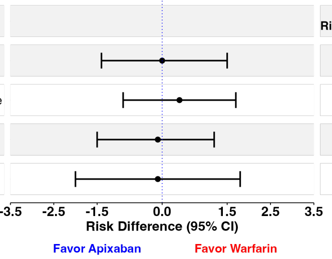


*Using stabilized weights from inverse probability treatment weighting (IPTW) and inverse probablity censroing weighting (IPCW); **Risk from IPT and IPC weighted cause-specifec cumulative incidence estimator; BMI:Body mass index; IPW: Inverse probability weighting; No.: Number of individuals in the analysis; RR:Risk ratio; SE: Systemic Embolism; 95% CI: 95% confidence interval estimated from non-parametric bootstrapping using 500 samples

**Figure S9.** Inverse probability weighted estimates of the 3-year BMI stratum-specific risks, risk differences per 100 people, and risk ratios of the comparative effectiveness for the total effect of apixaban versus warfarin in stroke/systemic embolism using a complete-case analysis and a hypothetical estimand (Estimand 8)

#### H.2 Analysis restricted to patients with BMI measurements in the last 3 years prior study entry

**Stroke/SE (IPW)***

| BMI groups | Apixaban (No.= 21,623) Events/No. | Risk** (95 %CI) | RR (95 %CI) | Warfarin (No.= 18,004) Events/No. | Risk** (95 %CI) | RR (95 %CI) | 36-month Risk Ratio (95% CI) |
|---|---|---|---|---|---|---|---|
| Overall | 594/21,623 | 4.4 (2.2,6.6) | | 683/18,004 | 3.5 (3.0,4.1) | | 1.25 (0.89,1.75) |
| Normal weight ≤24.9 kg/m2 | 198/5,837 | 4.8 (3.4,6.2) | 1.14 (0.76,1.70) | 197/4,158 | 4.2 (3.3,5.2) | Reference | 1.14 (0.76,1.70) |
| Overweight 25-29.9 kg/m2 | 227/7,525 | 4.1 (2.5,5.7) | 1.06 (0.76,1.48) | 240/6,212 | 2.8 (2.2,3.4) | 0.94 (0.58,1.53) | 1.48 (0.91,2.42) |
| Obese ≥30 kg/m2 | 169/8,261 | 4.5 (3.7,5.2) | 0.98 (0.60,1.59) | 246/7,634 | 4.0 (2.5,5.5) | 0.66 (0.44,0.97) | 1.12 (0.74,1.70) |

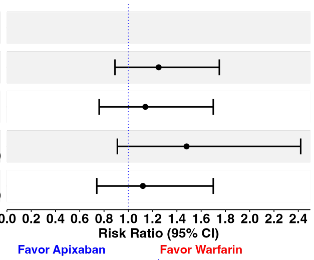


**Stroke/SE (IPW)***

| BMI groups | Events/No. | Risk** (95 %CI) | RD (95 %CI) | Events/No. | Risk** (95 %CI) | RD (95 %CI) | 36-month Risk difference (95% CI) |
|---|---|---|---|---|---|---|---|
| Overall | 594/21,623 | 4.4 (2.2,6.6) | | 683/18,004 | 3.5 (3.0,4.1) | | 0.9 (-1.4,3.2) |
| Normal weight ≤24.9 kg/m2 | 198/5,837 | 4.8 (3.4,6.2) | 0.6 (-1.1,2.3) | 197/4,158 | 4.2 (3.3,5.2) | Reference | 0.6 (-1.1,2.3) |
| Overweight 25-29.9 kg/m2 | 227/7,525 | 4.1 (2.5,5.7) | 0.3 (-1.0,1.5) | 240/6,212 | 2.8 (2.2,3.4) | -0.2 (-2.1,1.6) | 1.3 (-0.4,3.1) |
| Obese ≥30 kg/m2 | 169/8,261 | 4.5 (3.7,5.2) | -0.1 (-2.0,1.8) | 246/7,634 | 4.0 (2.5,5.5) | -1.4 (-2.6,-0.3) | 0.5 (-1.2,2.1) |

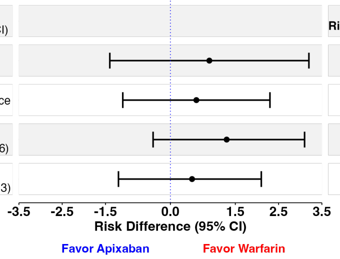


*Using stabilized weights from inverse probability treatment weighting (IPTW) and inverse probablity censroing weighting (IPCW); **Risk from IPT and IPC weighted cause-specifec cumulative incidence estimator; BMI:Body mass index; IPW: Inverse probability weighting; No.: Number of individuals in the analysis; RR:Risk ratio; SE: Systemic Embolism; 95% CI: 95% confidence interval estimated from non-parametric bootstrapping using 500 samples

**Figure S10.** Inverse probability weighted estimates of the 3-year BMI stratum-specific risks, risk differences per 100 people, and risk ratios of the comparative effectiveness for the total effect of apixaban versus warfarin in stroke/systemic embolism using a complete-case analysis for patients with available BMI measurements in the last 3 years prior study entry

| Major bleeding (IPW)* | Apixaban (No.= 21,623) | | | Warfarin (No.= 18,004) | | | |
|---|---|---|---|---|---|---|---|
| BMI groups | Events/No. | Risk** (95 %CI) | RR (95 %CI) | Events/No. | Risk** (95 %CI) | RR (95 %CI) | 36-month Risk Ratio (95% CI) |
| Overall | 1,486/21,623 | 7.4 (6.3,8.4) | | 2,189/18,004 | 9.3 (8.1,10.4) | | 0.79 (0.65,0.97) |
| Normal weight ≤24.9 kg/m2 | 428/5,837 | 10.4 (7.3,13.5) | 1.11 (0.75,1.66) | 517/4,158 | 9.4 (7.3,11.4) | Reference | 1.11 (0.75,1.66) |
| Overweight 25-29.9 kg/m2 | 518/7,525 | 6.8 (5.8,7.9) | 0.61 (0.45,0.82) | 728/6,212 | 10.0 (7.8,12.3) | 0.89 (0.65,1.22) | 0.68 (0.52,0.89) |
| Obese ≥30 kg/m2 | 540/8,261 | 5.7 (4.4,7.0) | 0.73 (0.55,0.96) | 944/7,634 | 8.4 (6.6,10.1) | 1.07 (0.78,1.48) | 0.68 (0.51,0.92) |

| Major bleeding (IPW)* | | | | | | | |
|---|---|---|---|---|---|---|---|
| BMI groups | Events/No. | Risk** (95 %CI) | RD (95 %CI) | Events/No. | Risk** (95 %CI) | RD (95 %CI) | 36-month Risk difference (95% CI) |
| Overall | 1,486/21,623 | 7.4 (6.3,8.4) | | 2,189/18,004 | 9.3 (8.1,10.4) | | -1.9 (-3.5,-0.3) |
| Normal weight ≤24.9 kg/m2 | 428/5,837 | 10.4 (7.3,13.5) | 1.1 (-2.7,4.8) | 517/4,158 | 9.4 (7.3,11.4) | Reference | 1.1 (-2.7,4.8) |
| Overweight 25-29.9 kg/m2 | 518/7,525 | 6.8 (5.8,7.9) | -3.6 (-6.0,-1.3) | 728/6,212 | 10.0 (7.8,12.3) | -1.0 (-3.6,1.6) | -3.2 (-5.7,-0.7) |
| Obese ≥30 kg/m2 | 540/8,261 | 5.7 (4.4,7.0) | -2.5 (-4.8,-0.2) | 944/7,634 | 8.4 (6.6,10.1) | 0.7 (-2.4,3.7) | -2.6 (-4.7,-0.5) |

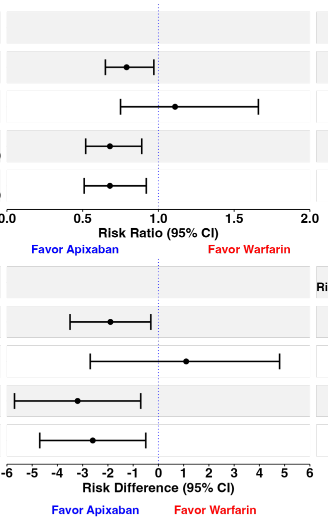


*Using stabilized weights from inverse probability treatment weighting (IPTW) and inverse probablity censroing weighting (IPCW); **Risk from IPT and IPC weighted cause-specifec cumulative incidence estimator; BMI:Body mass index; IPW: Inverse probability weighting; No.: Number of individuals in the analysis; RR:Risk ratio; 95% CI: 95% confidence interval estimated from non-parametric bootstrapping using 500 samples

**Figure S11.** Inverse probability weighted estimates of the 3-year BMI stratum-specific risks, risk differences per 100 people, and risk ratios of the comparative safety for the total effect of apixaban versus warfarin in major bleeding using a complete-case analysisfor patients with available BMI measurements in the last 3 years prior study entry

#### H.3 Direct effect with elimination of competing events

| Stroke/SE (IPW)* | Apixaban (No.= 27,747) | | | Warfarin (No.= 23,145) | | | |
|---|---|---|---|---|---|---|---|
| BMI groups | Events/No. | Risk** (95 %CI) | RR (95 %CI) | Events/No. | Risk** (95 %CI) | RR (95 %CI) | 36-month Risk Ratio (95% CI) |
| Overall | 719/27,747 | 4.3 (3.5,5.0) | | 842/23,145 | 3.7 (3.0,4.5) | | 1.14 (0.85,1.53) |
| Normal weight ≤24.9 kg/m2 | 236/7,669 | 5.3 (4.3,6.3) | 1.14 (0.82,1.60) | 243/5,691 | 4.7 (3.7,5.7) | Reference | 1.14 (0.82,1.60) |
| Overweight 25-29.9 kg/m2 | 292/10,280 | 4.6 (3.9,5.3) | 0.98 (0.72,1.33) | 318/8,561 | 4.4 (2.2,6.6) | 0.94 (0.59,1.52) | 1.04 (0.67,1.60) |
| Obese ≥30 kg/m2 | 191/9,798 | 4.2 (1.8,6.6) | 0.90 (0.53,1.51) | 281/8,893 | 3.4 (2.4,4.5) | 0.73 (0.47,1.13) | 1.23 (0.69,2.17) |

| Stroke/SE (IPW)* | | | | | | | |
|---|---|---|---|---|---|---|---|
| BMI groups | Events/No. | Risk** (95 %CI) | RD (95 %CI) | Events/No. | Risk** (95 %CI) | RD (95 %CI) | 36-month Risk difference (95% CI) |
| Overall | 719/27,747 | 4.3 (3.5,5.0) | | 842/23,145 | 3.7 (3.0,4.5) | | 0.5 (-0.5,1.6) |
| Normal weight ≤24.9 kg/m2 | 236/7,669 | 5.3 (4.3,6.3) | 0.7 (-0.8,2.1) | 243/5,691 | 4.7 (3.7,5.7) | Reference | 0.7 (-0.8,2.1) |
| Overweight 25-29.9 kg/m2 | 292/10,280 | 4.6 (3.9,5.3) | -0.1 (-1.3,1.1) | 318/8,561 | 4.4 (2.2,6.6) | -0.3 (-2.7,2.2) | 0.2 (-2.1,2.5) |
| Obese ≥30 kg/m2 | 191/9,798 | 4.2 (1.8,6.6) | -0.5 (-3.1,2.1) | 281/8,893 | 3.4 (2.4,4.5) | -1.3 (-2.7,0.2) | 0.8 (-1.8,3.4) |

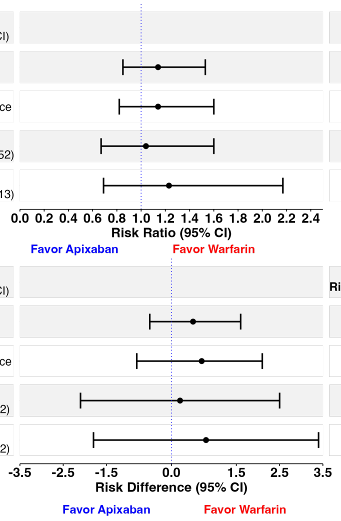


*Using stabilized weights from inverse probability treatment weighting (IPTW) and inverse probablity censroing weighting (IPCW); **Risk from IPT and IPC weighted Kaplan-Meier estimator; BMI:Body mass index; IPW: Inverse probability weighting; No.: Number of individuals in the analysis; RR:Risk ratio; SE: Systemic Embolism; 95% CI: 95% confidence interval estimated from non-parametric bootstrapping using 500 samples

**Figure S12.** Inverse probability weighted estimates of the 3-year BMI stratum-specific risks, risk differences per 100 people, and risk ratios of the comparative effectiveness for the direct effect of apixaban versus warfarin in stroke/systemic embolism using a complete-case analysis

| Major bleeding (IPW)* | Apixaban (No.= 27,747) | | | Warfarin (No.= 23,145) | | | 36-month |
|---|---|---|---|---|---|---|---|
| BMI groups | Events/No. | Risk** (95 %CI) | RR (95 %CI) | Events/No. | Risk** (95 %CI) | RR (95 %CI) | Risk Ratio (95% CI) |
| Overall | 1,205/23,145 | 7.2 (6.1,8.2) | | 1,852/23,145 | 9.0 (7.9,10.1) | | 0.80 (0.67,0.96) |
| Normal weight ≤24.9 kg/m2 | 359/7,669 | 11.0 (7.8,14.1) | 1.11 (0.74,1.65) | 469/5,691 | 9.9 (7.2,12.6) | Reference | 1.11 (0.74,1.65) |
| Overweight 25-29.9 kg/m2 | 445/10,280 | 6.4 (3.5,9.2) | 0.64 (0.45,0.92) | 659/8,561 | 9.0 (7.3,10.7) | 0.91 (0.65,1.27) | 0.71 (0.52,0.96) |
| Obese ≥30 kg/m2 | 401/9,798 | 6.9 (6.0,7.8) | 0.69 (0.52,0.93) | 724/8,893 | 10.4 (8.6,12.1) | 1.05 (0.76,1.44) | 0.66 (0.54,0.82) |

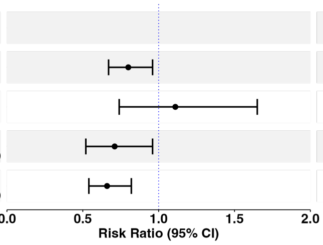


| Major bleeding (IPW)* | Apixaban | | | Warfarin | | | 36-month |
|---|---|---|---|---|---|---|---|
| BMI groups | Events/No. | Risk** (95 %CI) | RD (95 %CI) | Events/No. | Risk** (95 %CI) | RD (95 %CI) | Risk difference (95% CI) |
| Overall | 1,205/23,145 | 7.2 (6.1,8.2) | | 1,852/23,145 | 9.0 (7.9,10.1) | | -1.8 (-3.3,-0.3) |
| Normal weight ≤24.9 kg/m2 | 359/7,669 | 11.0 (7.8,14.1) | 1.1 (-3.1,5.2) | 469/5,691 | 9.9 (7.2,12.6) | Reference | 1.1 (-3.1,5.2) |
| Overweight 25-29.9 kg/m2 | 445/10,280 | 6.4 (3.5,9.2) | -3.6 (-7.5,0.4) | 659/8,561 | 9.0 (7.3,10.7) | -0.9 (-4.2,2.4) | -2.6 (-5.9,0.6) |
| Obese ≥30 kg/m2 | 401/9,798 | 6.9 (6.0,7.8) | -3.0 (-5.8,-0.2) | 724/8,893 | 10.4 (8.6,12.1) | 0.5 (-2.8,3.7) | -3.5 (-5.4,-1.6) |

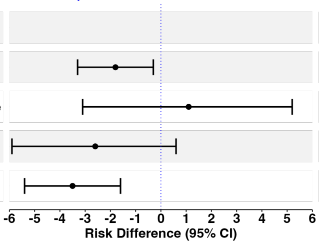


*Using stabilized weights from inverse probability treatment weighting (IPTW) and inverse probablity censroing weighting (IPCW); **Risk from IPT and IPC weighted Kaplan-Meier estimator; BMI:Body mass index; IPW: Inverse probability weighting; No.: Number of individuals in the analysis; RR:Risk ratio; 95% CI: 95% confidence interval estimated from non-parametric bootstrapping using 500 samples

**Figure S13.** Inverse probability weighted estimates of the 3-year BMI stratum-specific risks, risk differences per 100 people, and risk ratios of the comparative safety for the direct effect of apixaban versus warfarin in major bleeding using a complete-case analysis

#### H.4 Inverse probability weighted cox proportional hazard model

For the primary composite outcome, the hazard ratio (HR) (apixaban/warfarin) was 1.23 (0.98, 1.55) overall and 1.19 (0.89, 1.60) in normal weight group, 1.09 (0.86, 1.40) in the overweight group and 1.43 (0.83, 2.48) in the obese group.

For major bleeding, the hazard ratio (HR) (apixaban/warfarin) was 0.72 (0.62, 0.83) overall and 0.98 (0.72 , 1.32) in normal weight group, 0.60 (0.50, 0.72) in the overweight group and 0.67 (0.53, 0.83) in the obese group.

#### H.5 99% Truncated inverse probability weighting

| Stroke/SE (IPW)* | Apixaban (No.= 27,747) | | | Warfarin (No.= 23,145) | | | |
|---|---|---|---|---|---|---|---|
| BMI groups | Events/No. | Risk** (95 %CI) | RR (95 %CI) | Events/No. | Risk** (95 %CI) | RR (95 %CI) | 36-month Risk Ratio (95% CI) |
| Overall | 719/27,747 | 4.2 (3.9,4.6) | | 842/23,145 | 3.8 (3.3,4.3) | | 1.11 (0.94,1.31) |
| Normal weight ≤24.9 kg/m2 | 236/7,669 | 4.9 (4.2,5.7) | 1.17 (0.90,1.51) | 243/5,691 | 4.2 (3.4,5.1) | Reference | 1.17 (0.90,1.51) |
| Overweight 25-29.9 kg/m2 | 292/10,280 | 3.2 (2.6,3.7) | 1.12 (0.87,1.44) | 318/8,561 | 3.2 (2.4,3.9) | 0.97 (0.69,1.36) | 1.00 (0.73,1.38) |
| Obese ≥30 kg/m2 | 191/9,798 | 4.7 (4.1,5.4) | 0.75 (0.57,0.99) | 281/8,893 | 4.1 (3.1,5.1) | 0.75 (0.54,1.05) | 1.16 (0.87,1.53) |

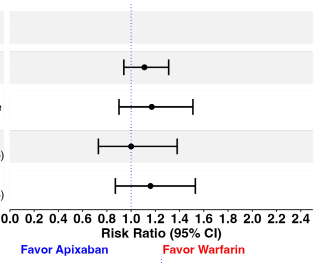


| Stroke/SE (IPW)* | | | | | | | |
|---|---|---|---|---|---|---|---|
| BMI groups | Events/No. | Risk** (95 %CI) | RD (95 %CI) | Events/No. | Risk** (95 %CI) | RD (95 %CI) | 36-month Risk difference (95% CI) |
| Overall | 719/27,747 | 4.2 (3.9,4.6) | | 842/23,145 | 3.8 (3.3,4.3) | | 0.4 (-0.2,1.0) |
| Normal weight ≤24.9 kg/m2 | 236/7,669 | 4.9 (4.2,5.7) | 0.7 (-0.4,1.8) | 243/5,691 | 4.2 (3.4,5.1) | Reference | 0.7 (-0.4,1.8) |
| Overweight 25-29.9 kg/m2 | 292/10,280 | 3.2 (2.6,3.7) | 0.5 (-0.5,1.5) | 318/8,561 | 3.2 (2.4,3.9) | -0.1 (-1.5,1.2) | 0.0 (-1.0,1.0) |
| Obese ≥30 kg/m2 | 191/9,798 | 4.7 (4.1,5.4) | -1.0 (-2.0,-0.1) | 281/8,893 | 4.1 (3.1,5.1) | -1.1 (-2.2,0.1) | 0.6 (-0.5,1.8) |

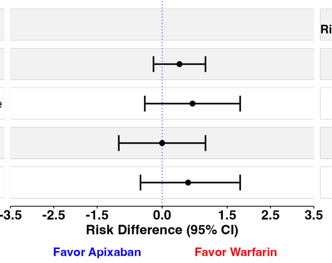


*Using stabilized weights from inverse probability treatment weighting (IPTW) and inverse probablity censroing weighting (IPCW); **Risk from IPT and IPC weighted cause-specifec cumulative incidence estimator; BMI:Body mass index; IPW: Inverse probability weighting; No.: Number of individuals in the analysis; RR:Risk ratio; SE: Systemic Embolism; 95% CI: 95% confidence interval estimated from non-parametric bootstrapping using 500 samples

**Figure S14.** 99%-truncated inverse probability weighted estimates of the 3-year BMI stratum-specific risks, risk differences per 100 people, and risk ratios of the comparative effectiveness for the total effect of apixaban versus warfarin in stroke/systemic embolism using a complete-case analysis

| Major bleeding (IPW)* | Apixaban (No.= 27,747) | | | Warfarin (No.= 23,145) | | | |
|---|---|---|---|---|---|---|---|
| BMI groups | Events/No. | Risk** (95 %CI) | RR (95 %CI) | Events/No. | Risk** (95 %CI) | RR (95 %CI) | 36-month Risk Ratio (95% CI) |
| Overall | 1,205/27,747 | 7.3 (6.8,7.8) | | 1,852/23,145 | 9.1 (8.3,9.8) | | 0.81 (0.73,0.90) |
| Normal weight ≤24.9 kg/m2 | 359/7,669 | 8.6 (7.4,9.8) | 0.96 (0.77,1.19) | 469/5,691 | 9.0 (7.4,10.5) | Reference | 0.96 (0.77,1.19) |
| Overweight 25-29.9 kg/m2 | 445/10,280 | 6.8 (6.0,7.5) | 0.75 (0.61,0.93) | 659/8,561 | 8.4 (7.2,9.6) | 0.94 (0.74,1.18) | 0.81 (0.67,0.97) |
| Obese ≥30 kg/m2 | 401/9,798 | 7.0 (6.2,7.7) | 0.77 (0.63,0.96) | 724/8,893 | 9.8 (8.5,11.1) | 1.09 (0.87,1.37) | 0.71 (0.60,0.84) |

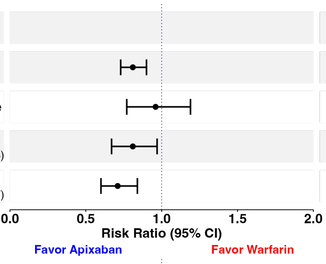


| Major bleeding (IPW)* | | | | | | | |
|---|---|---|---|---|---|---|---|
| BMI groups | Events/No. | Risk** (95 %CI) | RD (95 %CI) | Events/No. | Risk** (95 %CI) | RD (95 %CI) | 36-month Risk difference (95% CI) |
| Overall | 1,205/27,747 | 7.3 (6.8,7.8) | | 1,852/23,145 | 9.1 (8.3,9.8) | | -1.8 (-2.6,-0.9) |
| Normal weight ≤24.9 kg/m2 | 359/7,669 | 8.6 (7.4,9.8) | -0.4 (-2.2,1.5) | 469/5,691 | 9.0 (7.4,10.5) | Reference | -0.4 (-2.2,1.5) |
| Overweight 25-29.9 kg/m2 | 445/10,280 | 6.8 (6.0,7.5) | -2.2 (-3.9,-0.5) | 659/8,561 | 8.4 (7.2,9.6) | -0.6 (-2.5,1.4) | -1.6 (-3.0,-0.2) |
| Obese ≥30 kg/m2 | 401/9,798 | 7.0 (6.2,7.7) | -2.0 (-3.8,-0.3) | 724/8,893 | 9.8 (8.5,11.1) | 0.8 (-1.2,2.9) | -2.9 (-4.4,-1.4) |

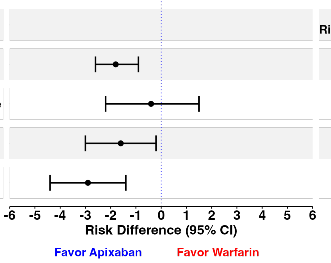


*Using stabilized weights from inverse probability treatment weighting (IPTW) and inverse probablity censroing weighting (IPCW); **Risk from IPT and IPC weighted cause-specifec cumulative incidence estimator; BMI:Body mass index; IPW: Inverse probability weighting; No.: Number of individuals in the analysis; RR:Risk ratio; 95% CI: 95% confidence interval estimated from non-parametric bootstrapping using 500 samples

**Figure S15.** 99%-truncated inverse probability weighted estimates of the 3-year BMI stratum-specific risks, risk differences per 100 people, and risk ratios of the comparative safety for the total effect of apixaban versus warfarin in major bleeding using a complete-case analysis
